# House modifications using insecticide treated screening of eave and window as vector control tool: evidence from a semi-field system in Tanzania and simulated epidemiological impact

**DOI:** 10.1101/2024.04.02.24305192

**Authors:** Olukayode G. Odufuwa, Richard J. Sheppard, Safina Ngonyani, Ahmadi Bakari Mpelepele, Dickson Kobe, Agathus Njohole, Jason Moore, Jastin Lusoli Lusoli, Joseph B. Muganga, Rune Bosselmann, Ole Skovmand, Zawadi Mageni Mboma, Emmanuel Mbuba, Rose Philipo, Jenny Stevenson, Ellie Sherrard-Smith, John Bradley, Sarah Jane Moore

## Abstract

**Background:** Despite extensive use of available vector control tools, the burden of malaria and dengue continues to increase throughout sub-Saharan Africa. Gaps in house structures, most especially in eaves and windows, allow vector entry and facilitate indoor vector biting and disease burden. Simple house modification tools that target these structures therefore have the potential to reduce human exposure to bites in the home. This study assessed the performance of Insecticide Treated Screening (ITS) comprising Eave Nets and Window Screens (ITENs & ITWS), incorporated with deltamethrin and piperonyl-butoxide (PBO) in Tanzania.

**Method:** A randomised Latin square (4 X 4) was conducted in four experimental huts built in a semi-field system (SFS). Each hut within each of the chambers of the SFS was covered with a large netting cage to allow recapture of mosquitoes inside and outside of the huts. Four treatment arms were evaluated: 1) new ITS; 2) 12-months naturally-aged ITS; 3) estimated 12 months field-used Olyset® Plus ITNs (Standard-of-Care in Tanzania), and; 4) no treatment. The study was performed for 32 nights using a minimum of 30 mosquitoes per strain per night, that is, a total of 120 (4 x 30) mosquitoes per hut per night. Four laboratory-reared strains were used: transmitters of malaria (*Anopheles arabiensis* and *An. funestus*) and dengue infection (*Aedes aegypti*) and those known for nuisance biting (*Culex quinquefasciatus*). Recaptured mosquitoes were assessed for mortality at 72 hours (M72), blood feeding and hut entry endpoints. A simulation exercise with a modified mechanistic model tracking *Plasmodium falciparum* malaria was used to illustrate the potential epidemiological impact from these products.

**Results:** New ITS induced higher M72 than field-used ITNs against all mosquito species tested [OR: 2.25 (95%CI: 1.65-3.06), p<0.0001], while M72 was similar between aged ITS and field-used ITNs [OR: 0.80 (95%CI: 0.59-1.08), p=0.141]. Both new, and aged ITS reduced more mosquito blood feeding and hut entry than field-used ITNs for all mosquito species tested (p<0.0001). Transmission model estimates indicate epidemiological impacts of ITS may supersede those of ITNs at the population level. The model results indicate that the potency of these impacts depends on assumed intervention percentage cover, durability and mosquito bionomics.

**Conclusions:** ITS is an efficacious tool for controlling vectors transmitting malaria, and dengue, and those known for nuisance biting in a semi-field setting. Given the intervention’s simplicity, it should be considered as an additional (or stand-alone) tool alongside behavioural change educational efforts to encourage the repurposing of old ITNs for house screening.

## Background

Insecticide treated nets (ITNs) and indoor residual spraying (IRS) are widely used and have contributed to the majority (>80%) [1] of the 2.1 billion malaria cases and 11.7 million malaria deaths averted globally in the period 2000–2022 [2]. ITNs are hung on the bedding space of sleepers, thereby providing an insecticidal barrier between the user and mosquitoes at night [3], while IRS is conducted on surfaces where mosquitoes rest [4]. The insecticides used in public health are generally contact toxicants that induce mortality; they may also disable mosquitoes, cause reduction in blood feeding, disorientation, or impede mosquito reproduction [5]. When these interventions are used at a large scale (protecting >80% of the community [6]) disease reduction among all community members is observed [7–12]. The widespread use of malaria control interventions also impacts other mosquitoes, including those that are of public health importance, such as those that transmit dengue infection [13] and community adherence is improved through the reduction in nuisance biting insects [14, 15]. These interventions remain a cornerstone for the control of mosquito vectors.

Despite over four decades of widespread use of ITNs and IRS, vector borne diseases, in particular malaria and dengue are increasing [2, 16, 17]. A possible explanation is the reduction in the impact of the core vector control tools. Evidence suggests that: 1) the insecticidal effectiveness is currently being hindered by increasing insecticide resistance among mosquitoes [18], caused by selection pressure from the wide use of the same insecticide class for the control of vector and agricultural pests [19–21]. Insecticide resistance needs to be managed to improve the effectiveness of the current classes of insecticide, but it is challenging to do this by increasing the dose or using a more potent class of ITNs because of the need to achieve critical safety standards [22] given the use of insecticide close to humans, technical considerations are required to incorporate or coat chemistries onto yarns [23], and the fact that ITNs are often washed. These reasons restrict the potential use of many ITN chemistries. 2) ITNs perform optimally when access (the proportion of households with ITNs for use by every two members) is sufficiently high [9], but recorded access to ITNs is currently low [4] despite the implementation of various strategies to improve it, such as behavioural change communication strategies and universal free ITNs distribution [24, 25]. Social hierarchies exist that render some people within households unprotected [26]. Furthermore, while the WHO - recommended ITNs are supposed to remain effective for a minimum of three years of operational use [10], however some data suggest that use-life is less than two years [27]. Mass campaigns delivering these ITNs usually occur at three year intervals leaving low access and use in the latter period prior to the next distribution [28]. 3) IRS is logistically challenging, requiring rigorous training and supervision of spray teams to ensure adequate coverage [4]. It also requires removal of all household belongings, which has been shown to affect its acceptability in the community [29]. It is also relatively costly [30] despite being extremely effective [31]. The residual efficacy of IRS in the field is usually below one year [32]. These limitations in core vector control tools call for additional low cost intervention that can be used for insecticide resistance management, lasts more than two years and requires minimal user compliance.

Reviews of studies on house screening [33–35] and a trial of a lethal house lure [35] conducted in a range of African settings report on the positive potential effect of house modification for malaria control. This makes sense as properly sealing housing structures such as eaves, windows, doors, ceilings, and traditional walls, or using iron sheet roofs or insecticide-incorporated or coated house structures may reduce indoor mosquito entry [33], and many mosquito species preferentially feed indoors at night when people are defenceless (asleep) and available [36]. Many houses in Africa, especially in the rural areas (where the majority of unimproved houses are found) have open eaves and windows that provide potential entry gaps but these structures are essential for indoor cooling and entry of natural light [37]. Addressing household entry by mosquitoes has the potential to protect all members of the household indoors and outside of sleeping hours [38, 39]. Although there are existing house modification tools, many of them are unsuitable for unimproved houses [34, 40] and are expensive to install. Simple closure of eaves increases indoor temperatures during the day, so is not acceptable in tropical regions [41].

The Insecticide Treated Screening (ITS) that include coverage of eaves (Insecticide Treated Eave Nets, ITENs) and windows (Insecticide Treated Window Screens, ITWS) [42] offer a potential solution to complement the core vector control tools. In a pilot study conducted in western Kenya [43], the netting used was incorporated with a pyrethroid insecticide and proved to be efficacious at reducing mosquito indoor densities, cost effective and acceptable to users. In order to control pyrethroid resistant mosquitoes, it is necessary to assess the efficacy of ITS incorporated with pyrethroids and Piperonyl butoxide (PBO) - a synergist used to restore susceptibility of mosquitoes to pyrethroids [44] due to the increasing resistance in mosquitoes to insecticide in sub-Saharan Africa (SSA) [18].

Experimental huts trials are used for assessing indoor intervention efficacy of ITNs and IRS [22, 45]. The entomological efficacy estimates are reliably assessed based on recaptured mosquitoes from inside the hut (or from within exit traps) [22]. In order to assess insecticidal housing modification including ITS, it is more appropriate to use a semi-field system (SFS) with huts built inside a netting cage [46, 47]. In this study, ITENs and ITWS were evaluated for their entomological efficacy against laboratory-reared pyrethroid-resistant malaria and dengue vectors, and nuisance biters in a SFS located in Tanzania, given that the tool has a potential of controlling populations of all these mosquitoes [48–50]. An established *Plasmodium falciparum* transmission model was adjusted to include the potential mechanism of action of screening houses with an insecticide treated fabric. We estimate malaria prevalence and incidence given the simulated implementation of ITNs, and ITS, and theorize on the potential epidemiological efficacy of these products.

## Methods

### Semi-field study location

The trial was conducted in the semi-field system (SFS) (Figure 1), of the Ifakara Health Institute (IHI) in Bagamoyo, Tanzania (6°25’42.4“S 38°53’01.9”E, Figure S1). The system measures 28.8 metres (m) X 21 m and is built on a raised concrete slab with a water filled ant channel. The walls were made of an untreated durable shade netting and the roof from polyethylene fabric (Fig 1a). The inside of the SFS was divided into four chambers, with a second large double-layered cotton netting cage (9 m width X 8.5 m length X 2.7 m height) suspended in each chamber with an Ifakara experimental hut [51] (Fig 1b). The inner cage (Fig 1c), white cloth lining in the hut and twelve resting buckets (Fig. 1d) were used to maximise mosquito recapture. One bucket was placed at each corner inside the experimental hut, one at each corner of the chamber and one at each side of the experimental hut (due to the large length and breadth of the large netting). White, heavy-duty cotton was used to cover the door and attached using Velcro (Fig 1e).

**Figure 1:**
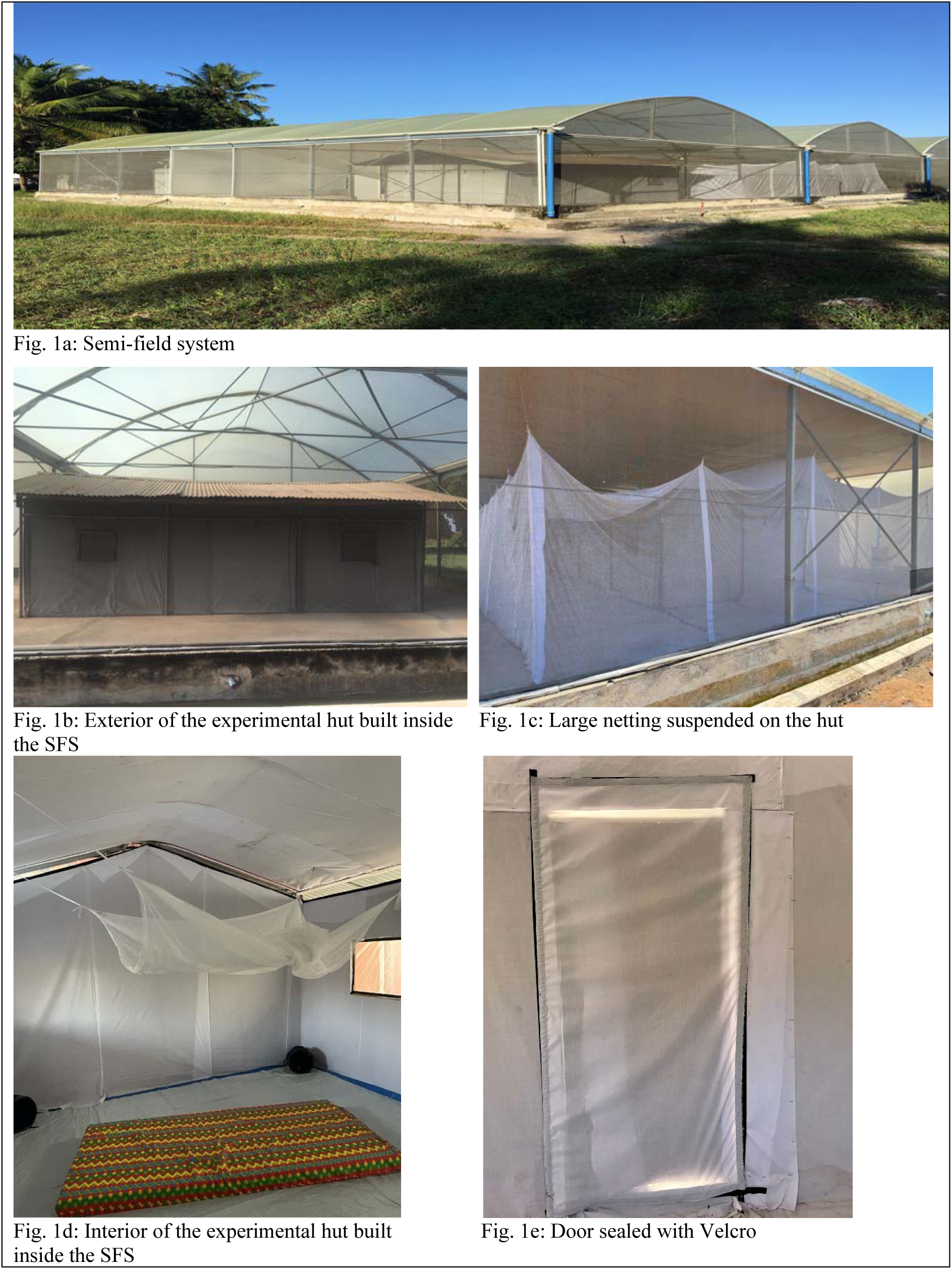
Semi-field system.

### Study design

A partially-randomised Latin square (4 X 4) design [52] was implemented. The aim was to compare the efficacy of new ITS (arm 1), and those that were naturally-aged for twelve months (arm 2), to aged (estimated one year in the field) Olyset® Plus ITNs (arm 3) and to a hut without any treatment (arm 4). Each arm was randomly allocated into each chamber on the first night of the test, and a sequential approach was employed thereafter; with nightly rotation of volunteers and rotation of arms at the end of every four nights. The test was conducted over thirty two experimental nights using four strains of laboratory-reared mosquitoes. Quality checks were conducted on net samples (25 cm X 25 cm) using WHO cone bioassay and High Performance Liquid Chromatography (HPLC) before and after the trial [22].

### Description of test items

The insecticidal netting for house modification [43] was a 0.152 mm diameter polyethylene yarn incorporated with 120 mg/m² deltamethrin and 540 mg/m² piperonyl butoxide (PBO). The nets were manufactured by Intelligent Insect Control, France. It was combined with PBO synergist as used for the control of mosquitoes with resistance to pyrethroids due to the upregulation of mixed function oxidases [44]. Olyset® Plus ITNs were selected as the comparator being the standard-of-care in Tanzania [53] and the first-in-class pyrethroid-PBO ITN [54], recommended based on a demonstration of its public health impact at the community level on malaria infection in epidemiological trials [55, 56]. The Olyset® Plus ITN is a 150-denier knitted monofilament polyethylene yarn incorporated with 20 g AI/kg (about 800 mg of AI/m^2^) permethrin and 10 g AI/kg (about 400 mg/m^2^) PBO, manufactured by Sumitomo Chemical Co., Ltd, Japan. Considering increasing numbers of households in the community without access to ITNs [57], it was necessary to have a hut without an intervention to assess the extent of exposure in local houses without any intervention. This “no treatment” arm also acts as the control comparator arm for the modelling simulations reported below.

### Description of study mosquitoes

Four laboratory-reared strains of mosquitoes were used in this study (Table S1). Three strains have resistance to pyrethroids that is fully or partially restored through pre-exposure to PBO: *An. arabiensis* and *An. funestus* are major malaria transmitter vectors in Tanzania [58, 59] and *Cx. quinquefasciatus* is notable for nuisance biting [60]. The fourth strain was pyrethroid-susceptible *Aedes. aegypti* that transmits dengue [61]. Mosquitoes released in the SFS chambers were nulliparous females, aged 5-8 days, sugar-starved for 6 hours before release. Mosquitoes exposed to cone bioassay for quality checks before and after the SFS were nulliparous sugar-fed and aged 3-5 days.

### Preparation of treatments for natural aging, washing and deliberate holing

In August 2021, ITS of approximately 0.5 m X 23 m were installed on the eaves (ITENs) and 1 x 0.7 m on each of the four windows (ITWS) of an animal house on the site of IHI in Bagamoyo using Velcro tape (Figure 2). They were left to naturally age until the end of September 2022.

**Figure 2:**
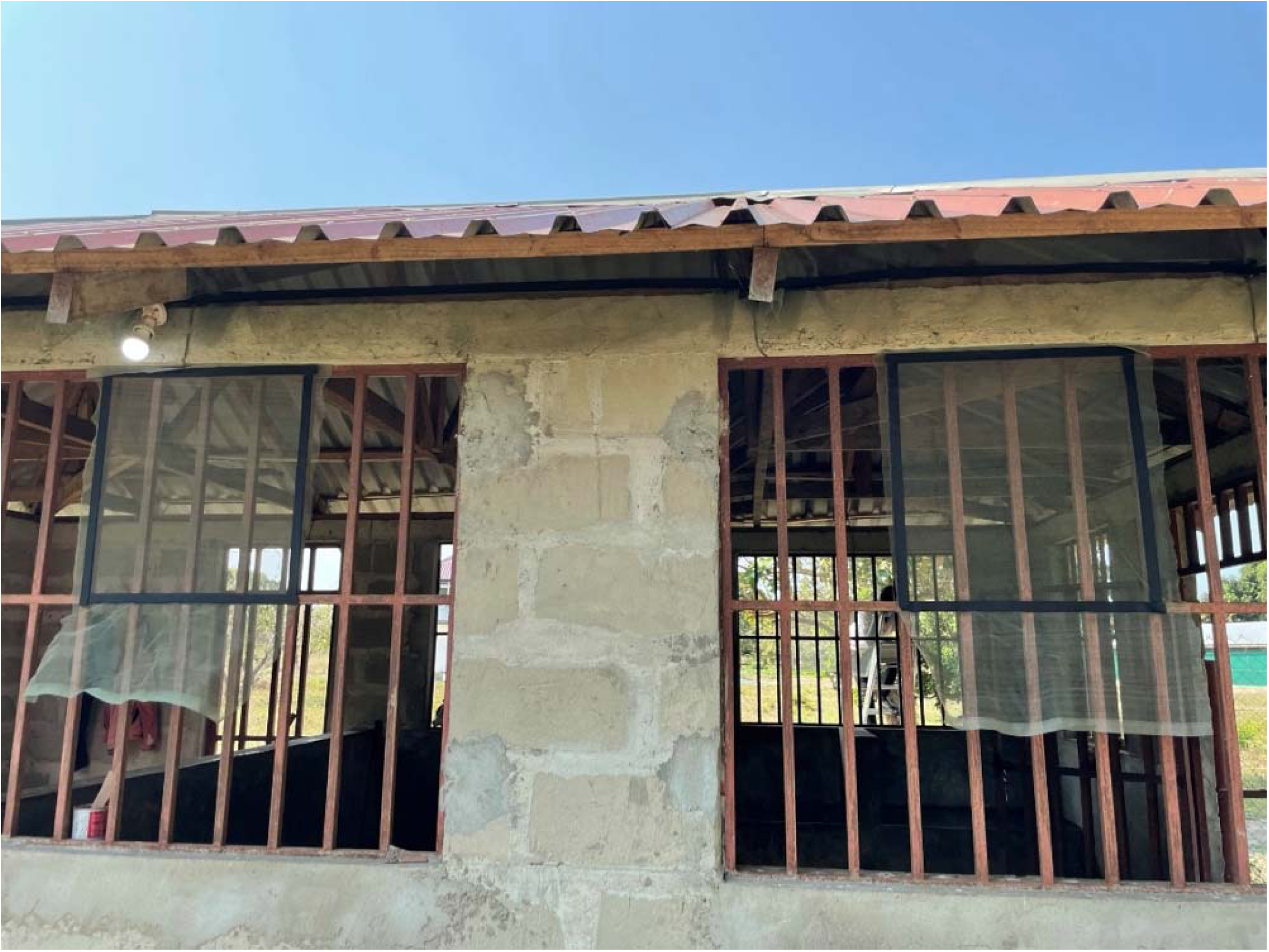
Natural aging of ITS.

On the same day of ITS’ installation, two new Olyset® Plus ITNs (1 for the experiment and 1 spare) were unwrapped from the packages and hung inside an experimental hut (Figure 3a), located 60 m from the animal house where ITS were installed. Olyset® Plus ITNs were periodically field-washed following WHO guidelines [62] in October and December 2021, as well as January, June, July and August 2022. This was based on average 6 annual washes reported from a community trial of Olyset® Plus ITNs in Tanzania [63]. Each of the ITNs was soap-washed once using 2 g/litre of soap (“Jamaa palm oil” soap flakes) and water-rinsed twice in an aluminium bowl (Figure 3b) filled with 10 litres of filtered well water (a maximum hardness of 5 dH). Each wash or rinse was standardised: stirring the net with a gloved hand at 20 rotations per minute for 3 minutes, soaking for 4 minutes and stirred for 3 minutes. Nets were dried horizontally in the shade where the conditions were no higher than 32°C and there was no direct sunlight (Figure 3c). After each wash and drying, nets were returned into the experimental hut for continued natural aging (Figure 3a).

Shortly before testing aged Olyset® Plus ITNs in the SFS, the nets were deliberately holed with scissors following estimated sizes and number of holes (Table 1) found on one year old Olyset® Plus ITNs tested under field conditions in Tanzania [63] (Figure 3d). The number of holes were equally divided into four sides of the nets.

**Figure 3:**
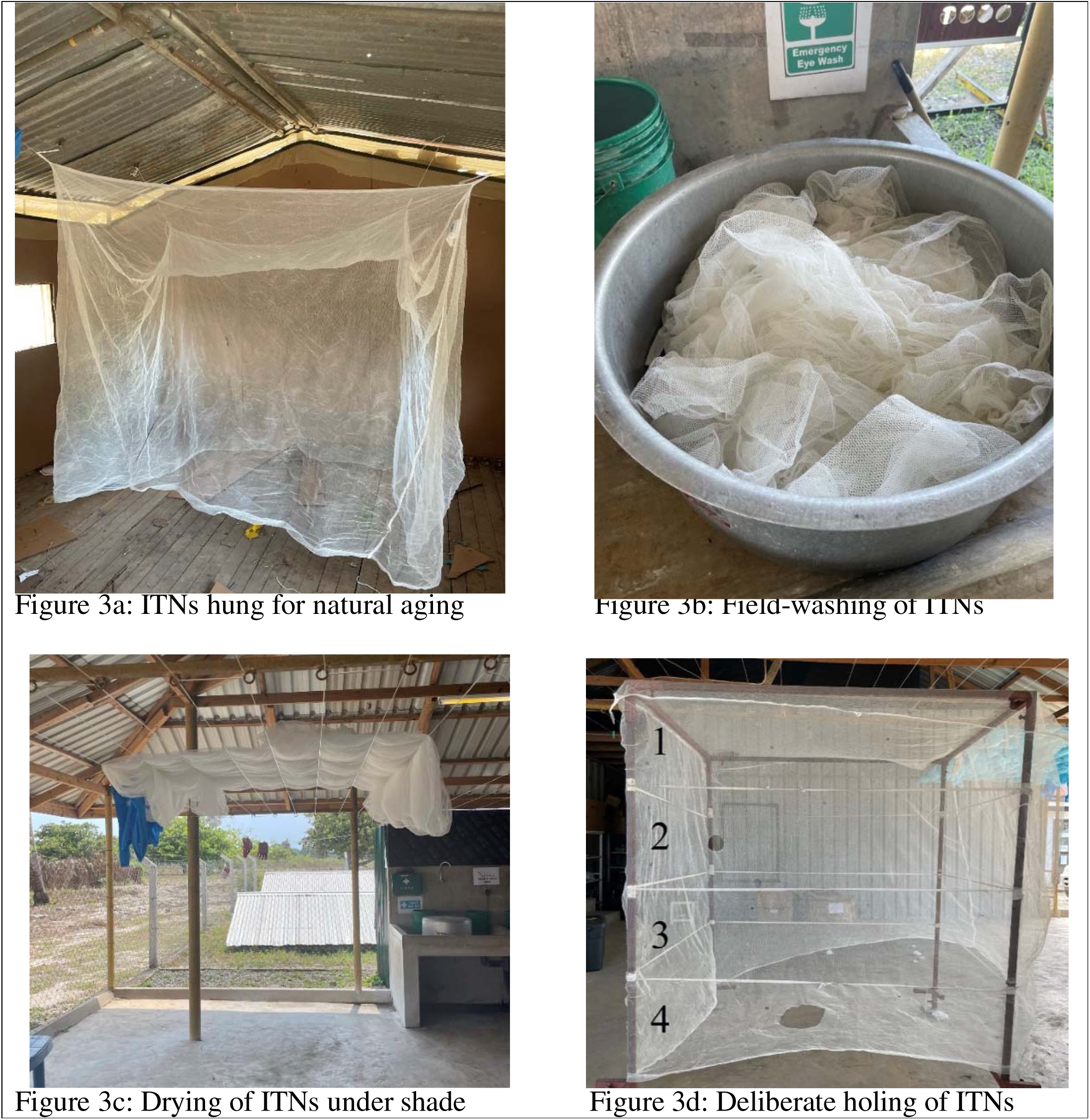
Natural aging of ITNs.

**Table 1:** Size and number of holes made on Olyset® Plus ITNs to simulate user condition.

| Size | Zone 1<br>(Upper) | Zone 2<br>(Under the<br>upper) | Zone 3<br>(Top of<br>bottom) | Zone 4<br>(Bottom) | Roof |
| --- | --- | --- | --- | --- | --- |
| Estimated<br>holes | 28 x size 1<br>(1.25cm) | 5 x size 1<br>(1.25cm)<br>1 x size 2<br>(6cm) | 14 x size 1<br>(1.25cm)<br>2 x size 2<br>(6cm) | 15 x size 1<br>(1.25cm)<br>4 x size 2<br>(6cm)<br>1 x size 3<br>(17.5cm) | 24 x size 1<br>(1.25cm)<br>5 x size 2<br>(6cm)<br>2 x size 3<br>(17.5cm) |

### SFS Experiment procedure

Testing in the SFS commenced in October 2022. Four male adults (≥18 years old) experienced in mosquito collection from the villages around IHI site of Bagamoyo were recruited and gave written informed consent for their participation in the study. Training on mosquito collection was provided to the volunteers after consent.

Each day of the experiment: 1) all chambers were cleaned with brooms that were fixed to individual chambers; 2) small towels were soaked in clean water before placement in each of the resting buckets; 3) study mosquitoes were allowed to acclimatize to the condition of the SFS for thirty minutes before release, and; 4) each of the volunteers were provided with four netted paper cups containing thirty mosquitoes per strain, mouth aspirators and mechanical Prokopack aspirators. At 18:00, after the inner net chamber was closed, each volunteer released the mosquitoes at a fixed point behind the experimental hut by gently opening the netted cups – leaving the freed mosquitoes to behave as naturally as possible, then volunteers entered the hut, shut the door and sealed the door with white cotton (Figure 1e). Each volunteer remained in the hut until 07:00 hours the next morning.

From 07:00 hours, mosquitoes were collected by volunteers using both mouth aspirators and a mechanical Prokopack, and additional checking was done by the supervisors. Volunteers recaptured mosquitoes in the experimental huts before recapturing those outside the hut within the chambers. The indoor collection was from the hut floor then resting buckets followed by the walls and roof. Recapture was conducted systematically from the bottom right corner of the hut, moving from the floor upwards in a clockwise direction, until all indoor mosquitoes were recaptured in netted paper cups. Outdoor capture was then systematically conducted in the inner netting chamber and resting buckets. After recapturing by volunteers, supervisors entered each chamber to verify that no mosquitoes are remaining in the chamber by gently shaking the cage netting or hitting the walls of the experimental hut. Cups were labelled by inside or outside of the hut, then transported to the sorting area. Mosquitoes were sorted and scored by location (inside or outside hut) as dead fed, dead unfed, alive fed and alive unfed and those living were held with access to 10% sugar solution for 72 hours in a temperature-controlled room to record M72.

In the first 4 days of the trial, a short questionnaire was administered to the participants that slept in the hut to record perceived adverse effects.

### SFS quality checks procedure

Before starting the SFS experiment, a sample (25 cm X 25 cm) was removed from the centre of each of the three rolls of insecticidal nets received to cover the eaves and windows of SFS. The pieces were removed after 100 cm piece was removed from each roll, given that the surface might have been exposed to unsuitable conditions during shipment. Three pieces were removed from one new Olyset® Plus ITNs at positions 5, 6 and 7 before testing and five samples were removed from aged Olyset® Plus ITNs from positions 1-5 of the 2013 WHO ITN testing guidelines [62]. After the SFS testing, samples (25 cm X 25 cm) were removed from the eaves and windows of the new and aged ITS (Figure S2) for testing.

All samples were tested against the 4 laboratory strains used in the SFS testing using cone bioassay with 4 replicates of 5 mosquitoes per sample (20 mosquitoes) following the 2013 WHO guidelines [62]. Mortality was observed until 24 hours for quality checks performed before SFS testing, and 72 hours for post SFS based on evidence of increased mortality at 72 hours for metabolic resistant mosquitoes [64]. Four samples (Figure S2, E1, E2, W1 and W2) were randomly selected for chemical analysis using the Collaborative International Pesticides Analytical Council (CIPAC) 331/LN/M/3 method.

### Data Management and analysis

Data was recorded on paper forms and double entered in Microsoft Excel (Microsoft). The two entries were merged to check if data was entered correctly before analysis was conducted in Stata 16 (Stata Corp, Texas, USA). Cleaning was done by checking daily balancing of treatments, chambers, volunteers and mosquito strains. Mosquitoes recaptured at each hour of assessment (12 and 72 hours) were verified against the number of mosquitoes released. Where discrepancies were found, paper forms were checked and corrections were made accordingly.

Descriptive statistics presented percentage arithmetic means with respective 95% confidence intervals of endpoints (M72 & blood feeding for SFS, and knockdown and M72 for cone bioassay). The median and interquartile range of the number of mosquitoes captured inside the hut was presented for deterrence endpoint in the SFS. Mortality in the untreated net was adjusted for treated nets using Abbott’s formula for cone bioassays because mortality in the untreated net was 5% [65].

Mixed effects binomial logistic regression was used to evaluate differences between arms for the proportion of M72 and blood feeding. A negative binomial mixed effects model was performed for the total number of mosquitoes recaptured indoor (deterrence). Hut position, sleeper, and day were factor fixed effects and observation (each row of data) was random effect to account for overall variation in the study.

### Transmission modelling to investigate potential epidemiological impact at the population level

The malaria simulation model [66] was adjusted to include a potential mechanism of action that replicates the installation of ITS. This *Plasmodium falciparum* transmission model has been described fully here [67, 68] and the house-screen updated code is publicly available (https://github.com/mrc-ide/malariasimulation; housing branch, accessed 29 March 2024).

Briefly, the human component of the model is an individual-based, stochastic transmission dynamics model. Humans are born as susceptible individuals inheriting maternal immunity that wanes through time. Mosquito biting rates are dependent on local human and mosquito interactions determined by assumptions on indoor and outdoor human activity, mosquito searching times and biting preferences. Biting rates are heterogeneous given population demographics, with fewer bites assumed on younger children given their smaller size. The risk of infection developing after an infectious bite, however, is assumed to decline with age due to naturally acquired immunity following continual exposure. A new infection may result in clinical illness or asymptomatic infection, and rates of these outcomes are impacted by individual immunity levels and may be prevented by interventions (e.g., implementing treatment strategies, vaccinations or other interventions). In the absence of treatment, individuals with clinical illness recover by moving through asymptomatic then sub-patently infected human states, corresponding with decreasing parasite prevalence. We only model death due to aging and do not explicitly model death due to disease.

The vector model is compartmental and deterministic, tracking different species of mosquito from eggs through to adults. Only adult females are tracked after emergence from pupae, and the sex ratio at emergence is assumed 50:50 following [69]. The mosquito carrying capacity is assumed to track rainfall patterns locally [68]. Adult mosquitoes seek hosts after emerging, can oviposit after successfully feeding and their mortality rate, prior to interventions are assumed as indicated in Table S2 for the species included in our simulation exercise (*An. arabiensis* and *An. funestus*). The proportion of bites taken on humans are estimated following [70, 71]. The probability that bites are attempted indoors or in bed in the absence of indoor interventions is estimated following [38]. Life expectancy, foraging and blood feeding rates are taken from Griffin et al. (2010) [72].

In the transmission model, the probability of a mosquito of a given species *v* biting host *i* during a single blood feeding attempt is defined as 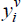 ; the probability of the mosquito biting and surviving the feeding attempt is 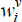, and the probability that she is repelled without feeding is 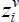 . Given these probabilities exclude natural vector mortality, with no protection these probabilities are 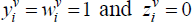 . Not all mosquitoes feed successfully when entering a house so 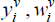 and 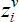 must take account the potential repeating behaviour of female mosquitoes foraging prior to introducing insecticides.

Following Le Menach et al. (2007) [73] and Griffin et al. (2010) [72], we add the potential impact from barriers to the model equations tracking the probable outcomes of a female vector feeding attempt through her oviposition cycle. During a single feeding attempt on either animals or humans, a mosquito will feed successfully with probability *W ^v^* such that:

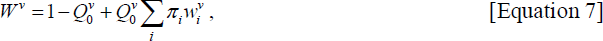

Where the last term in the equation is the probability that she will be repelled without feeding (*Z ^v^*):

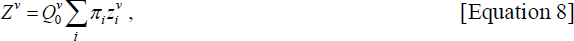

Here, 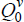 represents anthropophagy – and is defined in the model as the proportion of bites taken on humans by species *v* when interventions are absent (Table S2). Similarly, *π_i_* denotes the proportion of bites that person *i* receives, without interventions present. Protection due to house screening and ITNs, both depend on the proportion of bites that are taken when a person is protected by these interventions, and this depends on human sleeping patterns and mosquito activity. In the model, we define the proportion of bites taken indoors as *φ_I_* and the proportion of bites taken in bed as *φ_B._* Assumed values for these probabilities are shown in Table S2 for *An. arabiensis* and *An. funestus* mosquitoes.

When a mosquito attempts to enter a house, she can do so successfully (feeding and surviving, *s*), be killed (*d*) or be forced to repeat her search the following night (*r*). Here, we denote the success, death or repetition due to house screening as *s_h_, d_h_* and *r_h_* respectively following the notation from (Griffin et al 2010) [72], [a superscript *v* could be included for species but we leave this out for simplicity]. Similarly, *s_n_, d_n_* and *r_n_* show the corresponding outcomes with ITNs. Figure 4 demonstrates this pathway as depicted previously by Griffin et al. (2010) [72] with the addition of some form of housing barrier which induces a mortality effect (*d_h_*), and the removal of IRS impacts. The housing barrier is the first commodity that may be experienced by the indoor biting mosquito, where we assume that she may be repelled, killed or feed successfully. A mosquito that attempts to bite humans in bed may encounter an ITN where she may again be repelled, killed or feed successfully. If she survives (regardless of whether she was repelled or was able to feed), she must then exit the house passing the housing barrier a second time, where she may again be repelled, killed or exit the house. We assume that if she is repelled during her second encounter with the housing barrier, she moves back into the house space, where she will be killed eventually given she will encounter the screen on her attempt to exit.

The r*_h_*,, *d_h_* and r*_h_* terms are therefore included at both stages of the pathway, entering and exiting the building.

In the absence of house screening, both *r_h_* and *d_h_* are set to 0, and *s_h_* is set to 1, which means that mosquitoes cannot be trapped inside the house. Similarly, in the absence of ITNs, *r_n_* and *d_n_* are 0, while *s_n_* is 1.

**Figure 4.**
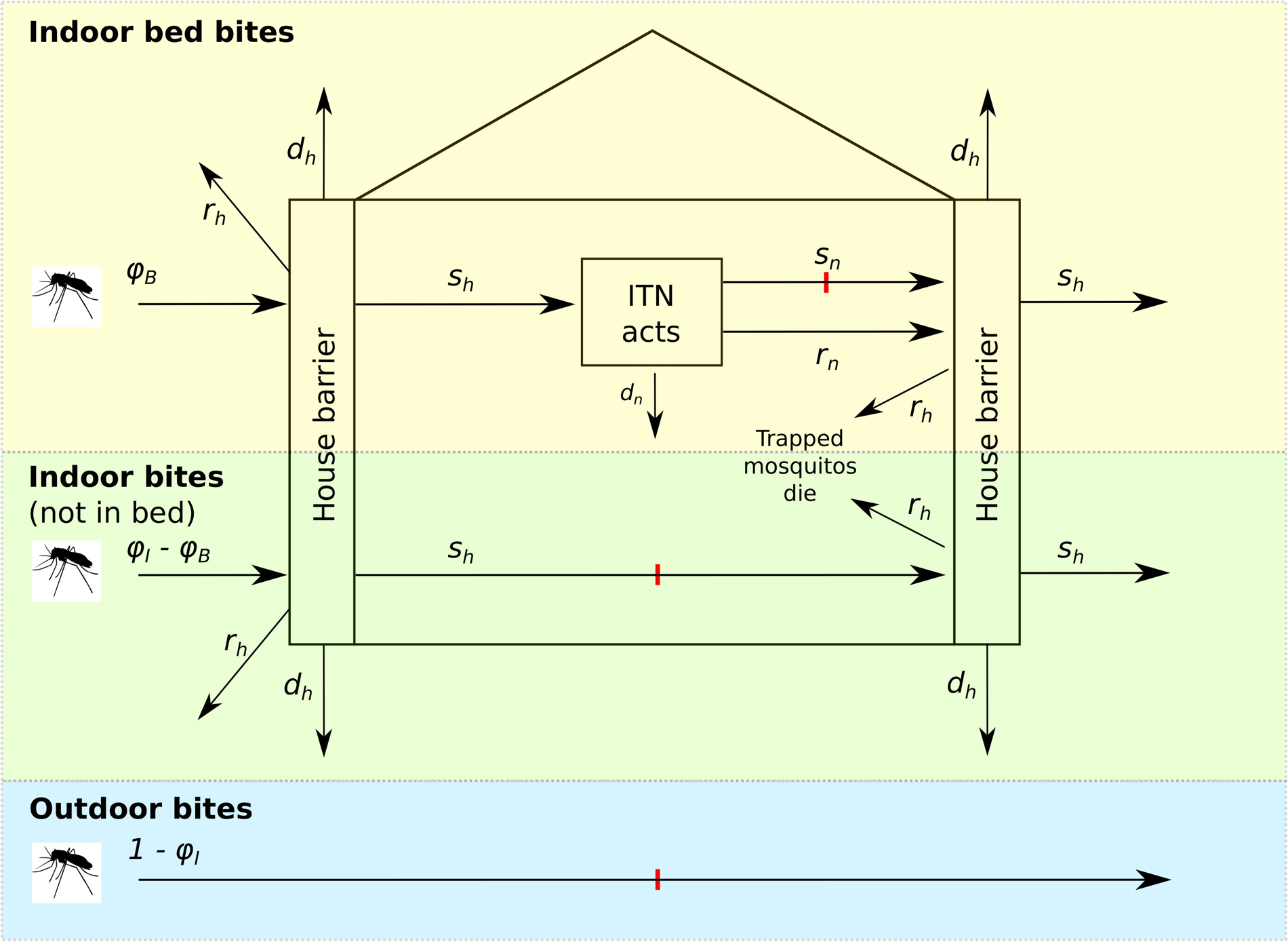
A schematic showing the modelled decision tree pathways for mosquito repetition, biting and survival, in a household with housing improvements (see subscript *h*) and where people may be sleeping under ITNs (see subscript *n*). We assume that a proportion of bites are taken outside (blue background; where there are no protective interventions), a proportion are attempted indoors but not on sleeping humans (green background; where they may encounter a housing barrier resulting in repetition *r_h_*, killing *d_h_* or survival and a successful bite *s_h_*) and a proportion are attempted on a sleeping human in bed (yellow background; where in addition to a housing barrier, they may additionally encounter an ITN, which again may result in repetition *r_n_*, killing before they are able to bite *d_n_*, or successful biting *s_n_*). Following a successful bite or repulsion by an ITN, a mosquito then must attempt to exit the house, passing again through the housing barrier, where she may again be repelled, killed or pass through safely having fed. This time, mosquitoes are repelled back into the house, where we make the assumption that they will eventually die. Bites occur at the positions on arrows marked with a red line. Symbol *φ_I_:* Proportion in mosquitoes attempting to bite humans indoors in the absence of interventions; *φ_B_:* Proportion of mosquitoes attempting to bite humans in bed in the absence of interventions.

Given this structure, it is possible to estimate the probability of a mosquito of a given species being repelled, biting or successfully feeding depending on the combination of interventions in action. We adjust the (Griffin et al 2010) [72] probabilities to include the effects from housing, (while removing the impact of IRS), and define the probability of surviving a successful feeding (*w_i_*) as:

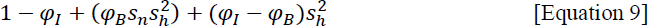

Similarly, we introduce the potential impacts from housing to the probability of biting (*y_i_*) such that:

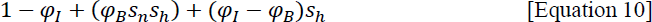

And the probability of repeating (*z_i_*) as:

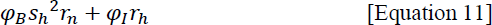

This mechanism allows us to set a fraction of a population to receive some form of protection from the housing adaptations, to simulate that the household changes reduce the entrance of mosquitoes by a defined quantity, and that some mosquitoes will be killed on contact with the screens.

In this contribution, the SFS data are used to quantify the effect from the ITS. In previous work, meta-analyses of experimental hut trials have been used to quantify the entomological impact of ITNs by taking the estimates for mosquitoes killed, fed or repelled (numbers in exit traps) in a given treated hut, and weighting these estimates by the deterrence estimate that is a comparison between mosquitoes caught within the untreated control hut and the treated hut [74] to generate parameter values for the model [67, 75]. Combining the data in this way allows an estimate of the model parameter values for *s_n0_*, *r_n0_*, and *d_n0_* that, respectively, represent the probabilities of mosquitoes successfully feeding, repeating or being killed when an ITN is present, works optimally (subscript 0 indicative of the net having been implemented on this day as a new product), and these values sum to 1 [67, 72, 75].

With the SFS data (see Supplementary data 1), we have estimates for the necessary model parameters for the ITN at 1 year, and for the ITS both new, and after 1 year. To choose representative model parameters, only the recaptured mosquitoes are considered. For each data point (daily count of mosquitoes given species and treatment), we determine the parameter estimates following the pathways in Figure 5. This means that, the mosquitoes recovered in the SFS data that are outside, fed and alive, out of the total recaptured, represent those who have passed into and out of the house in the screening treatment group. These are thus, *s ^2^*, so we must take the square-root to estimate *s_h_* for the model parameter. The corresponding proportion of mosquitoes who have been forced to repeat their feeding attempt *r_h_* can be estimated by the mosquitoes counted outside and alive, but unfed out of the total recaptured. Then, *d_h_ = 1 – s_h_ - r_h_*.

In the year-old ITN treatment, the proportion of mosquitoes that were recovered dead (either indoors or outdoors and either fed or unfed) represents the parameter *d_n,t= 365 days_*, where *t* can indicate optimal performance for newly applied netting (*t* = 0), or year-old performance (*t* = 365 days) as appropriate. The parameter *r_n,t= 365 days_* is calculated as the proportion of mosquitoes who are recaptured outside and alive out of the total recaptured during the SFS. The corresponding estimate for *s_n,t= 365_* is calculated as 1 – *r_n_,_t= 365 days_* – *d_n,t= 365 days_*.

Importantly, in the model, we need the estimates for these parameters at time 0, that is, the induced outcomes from products when they are newly introduced and working optimally. It is also necessary to estimate the durability of the products and make an assumption on how rapidly these impacts decay. This represents a critical unknown for both the pyrethroid PBO ITN and ITS products, and this is being addressed for the former in ongoing work [57]. Previously, a mean duration of mortality inducing impact has been estimated by Griffin et al. (2010) [69] from Mahama et al (2007) [73] for pyrethroid-only ITNs using washed ITNs, and the same logic applied for pyrethroid-PBO ITNs [75]. This assumes that the killing effect from ITNs will decrease at a constant rate *γ_n._*.

The impacts from implementing ITNs are projected to changes so that:

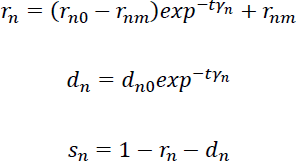

Applying these rates of decay to the data from the SFS, and back-calculating for the pyrethroid-PBO ITN entomological impact at time 0 returns higher mortality estimates than the meta-analysis (Table 2).

In the absence of further information for durability of the ITS, we explore a reasonable range in the mean duration of the product mortality inducing impact that would return mortality estimates after 365 days within the range of the SFS results. A minimum repellence impact is set for both ITNs (*r_mn_*) and screens (*r_mh_*). For ITNs, this estimate is assumed to be 0.24 (Griffin et al. 2010). For the screens, we cap this at 0.25 indicating a 25% reduction in entry given it is unlikely that the barrier will be lost even as insecticide potency wanes and it is assumed to be less likely that holes will accumulate as rapidly in the screen netting than the ITN netting. We mirror the structure for waning efficacy from ITNs into the mechanism of action for ITS such that:

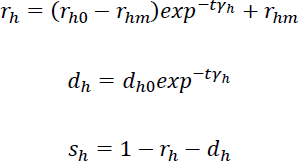

In addition, we include simulations that use pyrethroid-PBO ITN parameters developed via the statistical framework and validated against gold standard randomised control trial data for testing the epidemiological impact from these interventions [67]. In the review work preceding the validation exercise [74], associations between entomological impact and the level of phenotypic resistance in *Anopheles* populations were statistically determined allowing the resistance-level specific parameterisation of impacts on mosquitoes from ITNs within the transmission model. Values aligning with the resistance status of the species *An. arabiensis* and *An. funestus* tested within the SFS are used here for the model simulation exercise.

**Table 2.** Transmission model parameter ranges describing entomological impact from the pyrethroid-PBO ITN, and ITS as estimated from a systematic review of experimental hut trials [74] and modified for modelling and validated against RCTs [67], using the SFS data. Parameters shown are required to simulate the potential epidemiological impacts from these interventions.

| Parameter value and description |  | Parameter ranges simulated |  |
| --- | --- | --- | --- |
|  | Description | *An. arabiensis* outcomes: assuming 3 - 46 % of mosquitoes are killed on exposure to discriminating dose of pyrethroid at tube bioassay testing | *An. funestus* outcomes: (assuming 10 - 78 % of mosquitoes are killed on exposure to discriminating dose of pyrethroid at tube bioassay testing) |
| Pyrethroid PBO ITNs meta-analysis parameters |  | Median, 10<sup>th</sup> - 90<sup>th</sup> percentiles |  |
| $r_{n0}$ | Repellence outcome from pyrethroid-PBO ITN working optimally immediately after deployment | 0.500 (0.567 – 0.471) | 0.467 (0.551 – 0.444) |
| $d_{n0}$ | Mortality inducing effect from pyrethroid-PBO ITN working optimally immediately after deployment | 0.454 (0.261 – 0.504) | 0.500 (0.365 – 0.541) |
| $\gamma_n$ | Mean duration of mortality effect from pyrethroid-PBO ITN when working optimally | 808 (378 – 998) days | 995 (582 – 1277) days |
| Pyrethroid PBO ITNs semi-field system estimated parameters |  | Median, 25<sup>th</sup> – 75<sup>th</sup> percentiles |  |
| $r_{n0}$ | Repellence outcome from pyrethroid-PBO ITN working optimally immediately after deployment | 0.27 (0.46 – 0.24) | 0.35 (0.27 – 0.24) |
| $d_{n0}$ | Mortality inducing effect from pyrethroid-PBO ITN working optimally | 0.71 (0.51 – 0.75) | 0.72 (0.63 – 0.75) |
| $\gamma_n$ | Inferred mean duration of mortality impact from pyrethroid-PBO ITN when working optimally | 1392 (1016 – 1472) days | 1418 (1257 – 1472) days |
| ITS |  | Median, 25<sup>th</sup> – 75<sup>th</sup> percentiles |  |
| $r_{h0}$ | Repellence outcome from ITS working optimally immediately after deployment | 0.13 (0.04 – 0.25) | 0.04 (0.00 – 0.13) |
| $d_{h0}$ | Mortality inducing from ITS working optimally immediately after deployment (SFS data) | 0.73 (0.61 – 0.82) | 0.91 (0.82 – 0.95) |
| $\gamma_h$ | Mean duration of mortality effect from pyrethroid-PBO ITN when working optimally, inferred from SFS data | 774 (467 – 1795) days | 1284 (774 – 2101) days |

### Scenario analysis

We use the adjusted transmission model to simulate a set of theoretical intervention scenarios. To manage feasibility in the number of simulations run, and to specifically explore the comparison between pyrethroid-PBO ITNs and ITS, for all simulations, we use the median model parameters that have been fitted previously for malaria simulation [66, 72, 76, 77], except for the values indicated in Table 2. This is a limitation as we do not express full uncertainty, but this serves our purpose to illustrate potential impact from ITS, and comparable impact with pyrethroid-PBO ITNs. For the set up, we use a theoretical setting with malaria prevalence in children of 6 to 59 months of age reaching about 60% during the peak season, with a single transmission season profile. In all simulations, treatment of clinically ill patients is implemented so that 45% of people receive artemisinin combination therapy (ACT) when needed, a further 15% receive non-ACT treatment. It is assumed that 50% of mosquitoes have the *An. arabiensis-* like bionomics and 50% have the *An. funestus-*like bionomics (Table S2) and corresponding impacts from the ITN or ITS interventions (Table 2) given the range in resistance expressed during WHO tube bioassay testing (Table S1) as determined by Nash et al. (2021) [74] and validated in Sherrard-Smith et al. (2022) [67]. Adherence to ITN use is also simulated to wane over time so that by 3 years, half as many people are using the original net. This is a reasonable assumption that net use wanes over time [27]. Each simulation is run for 15 years, the first 6 years generate a relative equilibrium before we switch on comparative scenarios (in Figure 6, this is year 0). Pyrethroid-PBO ITNs are implemented every 3 years from the start, depending on the scenario explored, ITS is also implemented on 3-year cycles from year 0 (Figure 6). It is assumed for both that the interventions are deployed overnight and to the target proportion of the population, who immediately replace old nets with the newly distributed ones and gain the benefit from their impact. In reality, it will take longer to provide a community with ITS, but the longer-term gains should level out for this theoretical exploration of epidemiological impacts.

To consider the potential impact, of each intervention class (pyrethroid-PBO ITNs or insecticide treated house screening), a counterfactual simulation is also run where ITNs are distributed in year -6 and -3 but then no longer deployed. Older nets may still circulate in the population but will have a far lower efficacy given insecticide would have waned and adherence to use fallen to a low level. To estimate reductions in prevalence, the average prevalence in children of 6 to 59 months of age, 5-15 years and for all-ages, over years 0-3 for the treatment simulation (*C_T_*) are compared to the corresponding estimate from the counterfactual simulation (*C_C_*). To estimate cases averted, the same age groups are considered across 3 years to generate an estimated number of cases averted per 1,000 population. That is:

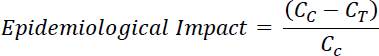

Where *C* can indicate prevalence or cases as required.

The pyrethroid-PBO ITNs – parameterised using the meta-analysis approach [67, 74] or using the directly comparable approach by generating parameters from the SFS data – and ITS products are simulated as stand-alone or combined interventions (Figure 6). Code is provided as Supplementary Material 2.

Next, to consider the potential of this novel product, a simple sensitivity analysis is performed on the population ITS coverage and product durability (the mean duration of the mortality inducing impact). The counterfactual is the use of pyrethroid-PBO ITNs, parameterised as per the meta-analysis [67, 74], distributed every 3 years to 60% of the population on deployment. Here, we simulate the median estimated impact from the ITS given increasing cover from 0 to 80% of the community, and for a product that has a mean duration of 1 year through to 5 years.

## Results

### Experimental validity

Throughout the whole year of natural aging in the experimental hut, the median temperature was 27.2°C [Interquartile range (IQR): 24.3-32.6°C) and relative humidity was 82.0% (IQR: 60.8 – 93.1%).

Nightly (6:00 PM to 7:00 AM), the median temperature and relative humidity in chambers outside of the experimental huts was 26.4°C (IQR: 25.2 – 27.2) and 91.4% (IQR: 85.5 – 97.0), respectively. The median indoor temperature of huts installed with ITS was 25.2 °C C (22.6 – 26.7), and this was similar to huts with Olyset® Plus ITNs 25.0 °C C (21.6 – 26.2), and no treatment huts 25.2 °C C (24.0-26.3), indicating that ITS did not increase indoor temperature.

Recaptured mosquitoes from the chambers of the SFS were held at median temperature of 25.6°C C (IQR: 25.4 – 25.8) and relative humidity of 78.3% (71.9 – 82.4) to assess M72, meeting WHO thresholds for temperature (27±2 ^O^C) and relative humidity (80±20%) for laboratory testing [22] .

Overall, 85% (13,370 of 15,722) of the mosquitoes released were recaptured in this study: 85% (3,300 of 3,883) *An. arabiensis*, 80% (3,186 of 3,940) *An. funestus*, 87% (3,427 of 3,929) *Cx. quinquefasciatus* and 87% (3,457 of 3,970) *Ae. aegypti* mosquitoes. The proportion of overall mosquitoes recaptured varied slightly among the arms with more mosquitoes recaptured in the untreated arm (93%) than the aged Olyset® Plus ITNs (81%), new ITS (78%) and aged ITS (87%) (Table S3).

### ITS has potential for providing community effect against mosquito vectors

ITS induced mortality to mosquitoes measured at 72 hours. Overall (all strains combined), the M72 among mosquitoes exposed to the aged ITS were similar to those exposed to the aged Olyset® Plus ITNs [51% vs 56%, Odds Ratio (OR): 0.80 (95% Confidence Interval: 0.59 – 1.08), p-value = 0.141], whereas the M72 was significantly higher for new ITS [68% vs 55%, OR: 2.25 (95% CI: 1.65 – 3.06), p-value <0.0001] in comparison to aged Olyset® Plus ITNs (Table 3 and Figure 5).

There was variation in the M72 observed within the strains (Table S4). While *An. arabiensis*, *Cx. quinquefasciatus* and *Ae. aegypti* mosquitoes tested against aged ITS died at similar proportions to those exposed to aged Olyset® Plus ITNs (p>0.05), a significantly lower M72 was observed for *An. funestus* mosquitoes exposed to the aged ITS arm in comparison to the aged Olyset® Plus ITNs [66% vs 79%, OR: 0.44 (95% CI: 0.27 – 0.72), p=0.001]. For the new ITS, significantly higher proportions of *An. arabiensis*, *An. funestus* and *Ae. aegypti* mosquitoes died at 72 hours after exposure to ITS compared to those exposed to the aged Olyset® plus ITNs, however no significant difference was found for *Cx. quinquefasciatus* mosquitoes. All treatments had higher mortality than the untreated arm (Table S4).

**Table 3:**
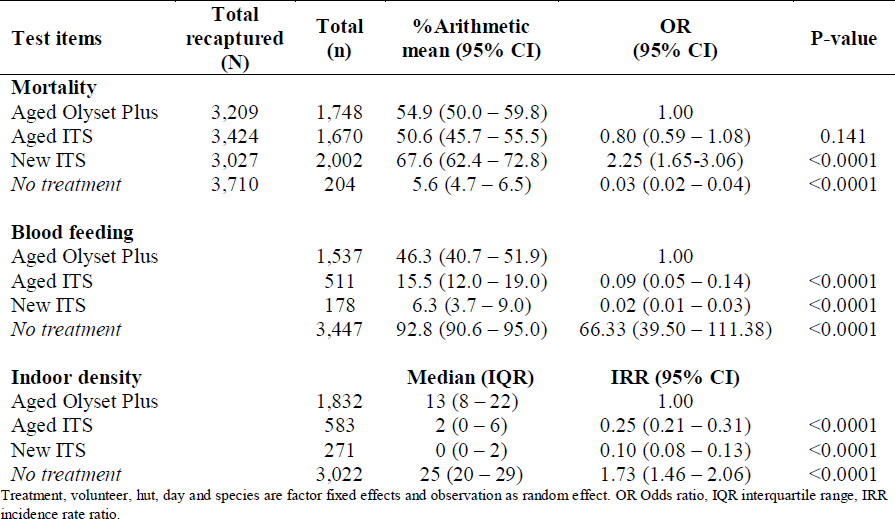
Mortality at 72 hours, blood feeding and deterrence of mosquitoes exposed to Insecticide Treated Eave Nets and Insecticide Treated Window Screens (Insecticide Treated Screening, ITS) compared to a standard of care pyrethroid-PBO net, Olyset Plus in the semi-field system. All mosquito strains combined.

**Figure 5:**
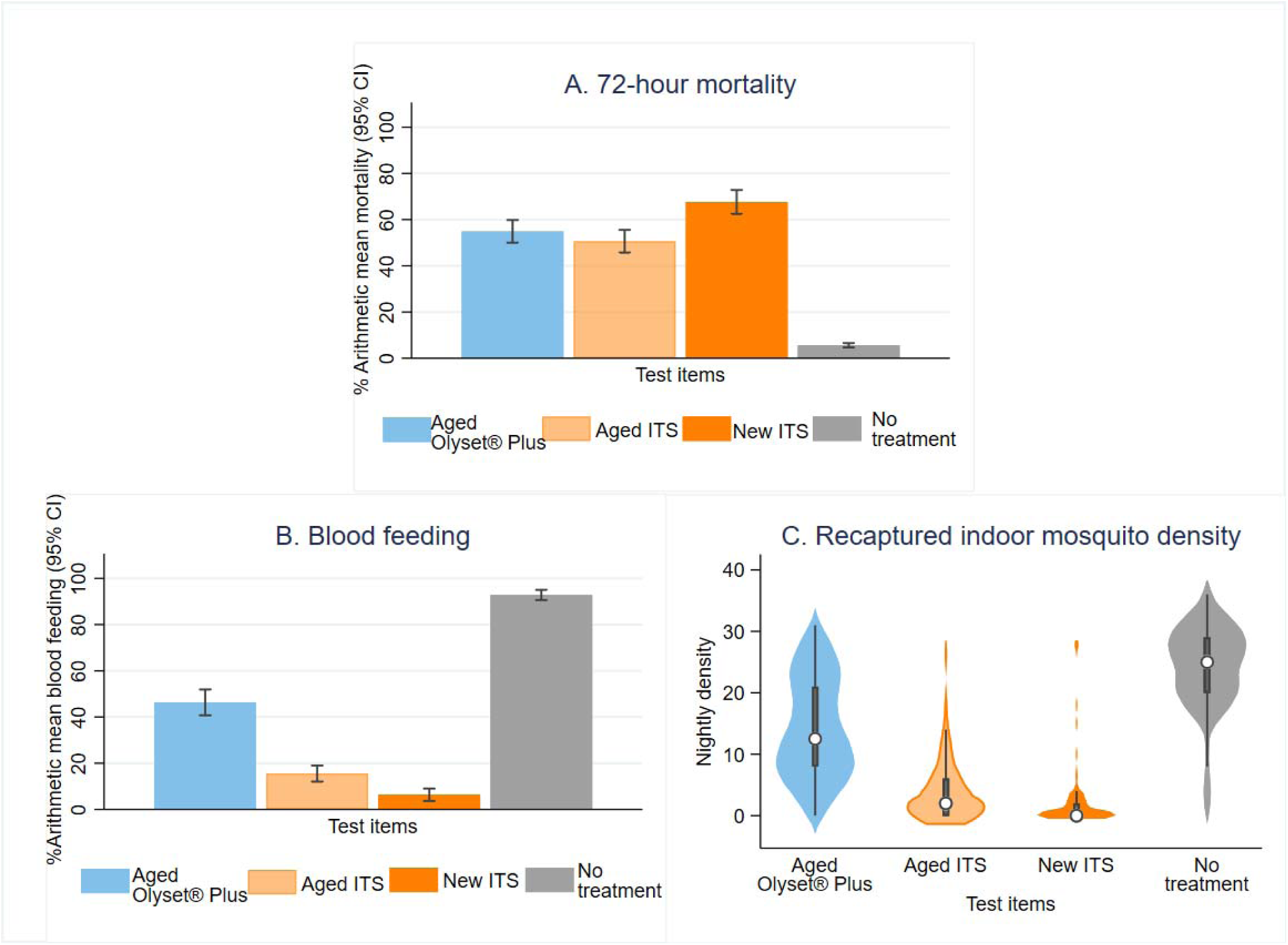
The proportion of mortality (A) and blood feeding (B), and median indoor density of overall mosquitoes (pyrethroid-resistant *An. arabiensis*, *An. funestus, Cx. quinquefasciatus* and susceptible *Ae. aegypti*) recaptured after exposure to experimental huts installed with ITS against the standard of care: Olyset® Plus ITN in the semi-field system in Tanzania.

### ITS provides personal protection

Of all the mosquitoes recaptured, significantly fewer blood-fed mosquitoes were recovered in the experimental huts installed with ITS that were new [6%, OR: 0.02 (95% CI: 0.01 – 0.03), p-value <0.0001] or aged [16%, OR: 0.09 (95% CI: 0.05 – 0.14), p-value <0.0001] compared to huts installed with aged Olyset® Plus ITNs (46%), Table 3 and Figure 5. Almost all the strains had significantly lower proportions of blood feeding in the huts installed with ITS (p<0.0001), with the exception for *An. funestus* mosquitoes, that blood-fed in similar proportions in the huts installed with aged ITS to huts with aged Olyset® Plus ITNs [OR 0.63 (95% CI: 0.32 – 1.24), p=0.182] (Table S5).

### ITS prevents mosquito indoor entry

Installation of ITS in the experimental huts significantly reduced entry of all mosquito strains tested in this study, both new [median 0, IRR: 0.10 (95% CI: 0.08 – 0.13), p-value <0.0001] and in aged condition [median 2, Incidence Rate Ratio (IRR): 0.25 (95% CI: 0.21 – 0.31), p-value <0.0001] compared to aged Olyset® Plus ITNs with a median 13 mosquitoes caught indoors (Table 3, Figure 5). Reduction in indoor densities was significantly higher for new and aged ITS than aged Olyset® Plus ITNs among all the strains tested (Table S6, Figure 5).

### Perceived adverse effects

None of the volunteers withdrew from the study due to any perceived adverse effect. Although, sneezing was the common symptom reported by the participants of the trial (Table S7).

### Quality checks Cone bioassay and Chemical analysis

Median temperature was 26.0°C C (IQR: 25.9 – 27.0) and relative humidity was 69.8% (52.4 – 76.4) during the implementation of cone bioassays, meeting acceptable, recommended bioassay conditions. Cone bioassays conducted on three pieces (measuring 25 cm X 25 cm) of ITS before the implementation of the SFS revealed complete knockdown (100%) after sixty minutes of exposure and minimum mortality of 73% at 24 hours against *An. arabiensis*, *An. funestus Ae. aegypti* mosquitoes (Table S8). After the SFS, the proportion of knockdown and mortality from new and aged ITS and Olyset® Plus reduced drastically (Table S8). While the chemical analysis revealed that low deltamethrin content (27%) and PBO synergist (8%) were retained after aging of ITS (Table S9).

### Simulated Epidemiological potential of ITS

The SFS data indicated that the pyrethroid-PBO ITNs in the tested system (Supplementary Figure 3) were working better than the average parameter estimates for the product derived from a meta-analysis review of experimental huts [67, 74] Table 2. This translates to an improved epidemiological impact simulated in the transmission model for the SFS derived parameters, where the SFS derived parameters are at the upper bound of the meta-analysis derived parameter uncertainty shown in Figure 6A. The reduction in mean prevalence in children of 6 to 59 months of age over 3 years, relative to a scenario simulating the cessation of mass ITNs distribution previously achieving 60% use by the population at implementation, that is achieved by the continuation of the pyrethroid PBO ITN campaign is 30.2 (21.7– 30.9) % for the SFS parameterisation, and 15.2 (3.3 – 21.3) % for the meta-analysis parameters. The additional implementation (given SFS parameters) of the ITS product to cover 60% of the population in a comparable scenario reduces prevalence in the same age cohort by 77 (68 – 81) % (and this is 71.4 (62.5 - 78.0) % for the meta-analysis pyrethroid-PBO ITN parameters together with ITS). Used alone, the modelled impact from the ITS remains impressive, achieving a reduction in prevalence among children of 6 to 59 months of 65.2% (51.2 – 72.1) relative to no further ITN campaigns from year 0.

The sensitivity analysis shows that increasing the durability of the mortality inducing impact and increasing the cover of people within the population who are benefiting from the product is beneficial (Figure 6B and C). Even when the mean duration of impact on mortality is low, between 1 and 2 years, achieving screening for 60% of the population may be able to reduce mean prevalence in children of 6 to 59 months of age over 3 years by 60% compared to no ITNs (Figure 6B), and all age clinical incidence by over 80% (Figure 6C).

**Figure 6.**
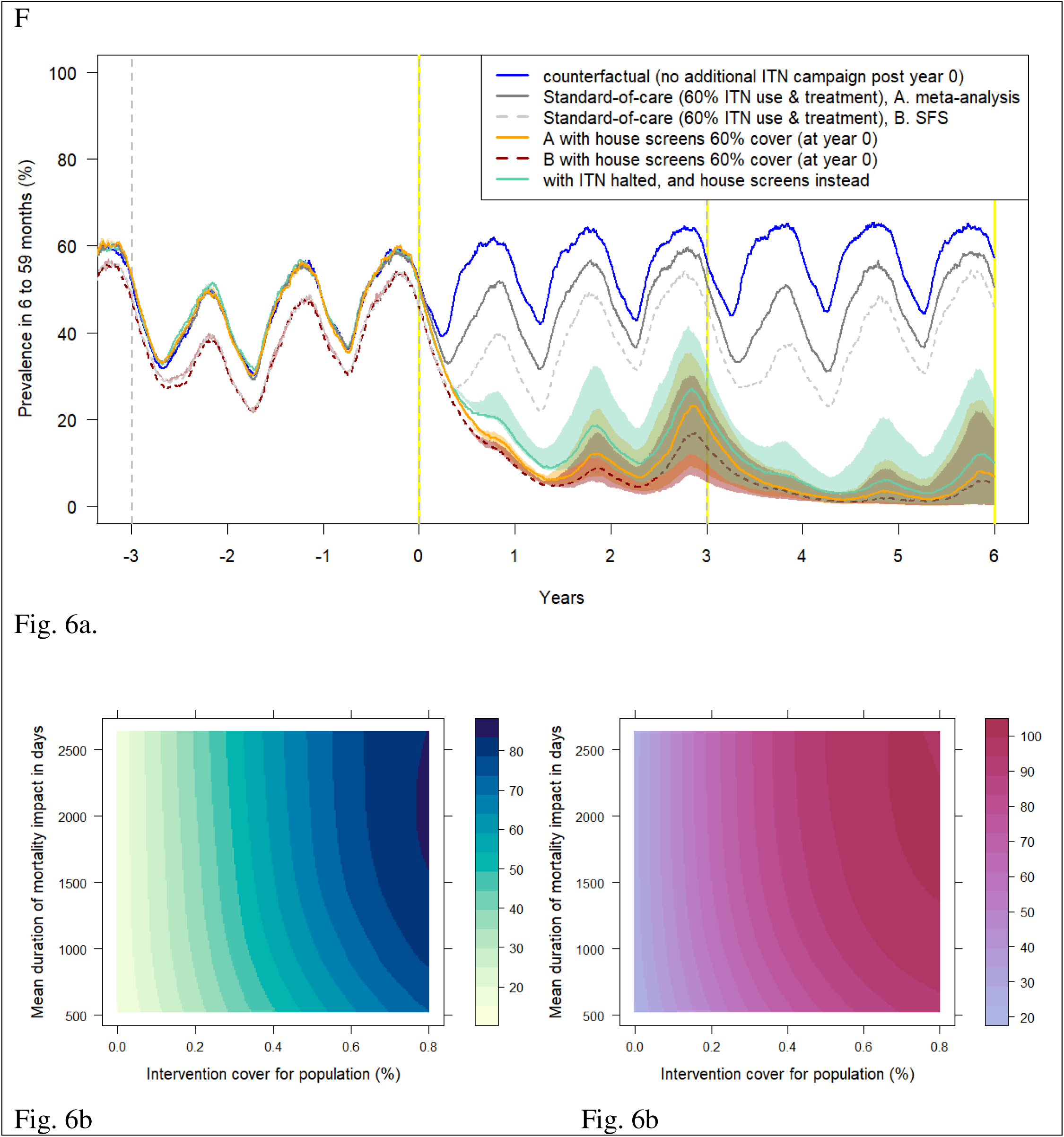
A transmission simulation exercise to explore potential epidemiological impact from the ITS. A) A theoretical set of scenarios showing the microscopy measured *falciparum* parasite prevalence in children of 6 to 59 months of age over time given implementation of ITNs, and ITS, covering 60% of people on implementation. ITN distribution occurs every 3 years (vertical grey dashed lines). Adherence to use wanes with a mean duration of 3 years, and insecticide durability wanes according to estimates from Sherrard-Smith et al 2022 [67], grey solid line) or the SFS data (grey dashed line). The effect of removing this ITN intervention from year 0 onward is indicated by the blue line (where the corresponding incidence outputs from this simulation are used as the respective counterfactuals for panels B and C). The impact ITS used in addition to ITNs is shown by the solid orange line (with meta-analysis parameters for ITNs), and red line (with SFS data parameters for ITNs). The impact from ITS as an alternative to ITNs is shown as the green line. Uncertainty that is due to the upper and lower estimates shown in Figure 6A are demonstrated by the corresponding polygons. All other model parameters are matched between runs. B) The estimated relative reduction in mean prevalence in children of 6 to 59 months of age given deployment of an ITS with specified population cover and mean duration of mortality impact on mosquitoes, compared to the counterfactual simulation, over the 3 years (time 0 to 3 years in panel A). C) The percentage reduction in all-age clinical incidence against the same counterfactual.

## Discussion

The most impactful mode of action of vector control products is the ability to induce mortality to mosquito vectors [78, 79], so that it can sufficiently reduce their population size and age to reduce overall burden of disease among users and non-users of the intervention. The experimental huts installed with ITS killed more than half of the pyrethroid-resistant mosquitoes released due to the combination of deltamethrin and PBO synergist incorporated into it [44]. The corresponding modelling exercise offers support that as an additional vector control tool, it is capable of impacting malaria prevalence. The concept of shielding entry points with insecticidal barriers has been shown to be successful in a trial of eave tubes that incorporated an insecticide with mortality as the primary mode of action [35]. Given the increasing resistance to pyrethroids in Africa [18] and the reduced application of IRS [80] for insecticide resistance management the use of non-pyrethroids on ITS would be beneficial [81]. The use of ITS may offer an alternative delivery method for insecticides that are water soluble or with a toxicity profile not suitable for ITNs [5], thus increasing the possible active ingredients available for the control of vector borne diseases.

A similar proportion of mosquito mortality was observed between huts installed with aged ITS (51%) and aged Olyset® Plus ITNs (55%). Confirmatory cone bioassay and chemical testing also indicated substantial loss of bio efficacy and loss of active ingredient during the process of aging. ITS was exposed to environmental conditions including temperature, wind, and dirt. ITWS was exposed to Ultraviolet (UV) light through the sun light. Studies have shown that degradation of insecticide in nets is principally due to UV [82], and the long term exposure of ITWS to sun light in the animal house might have contributed to the degradation of the insecticide and synergist incorporated within it. While it was not clear if ITENs was exposed to direct sunlight, evaporation might be a major reason for insecticidal loss of the ITENs. The Olyset® Plus ITN used in this experiment was not exposed to direct sun light as it was hung inside the experimental system where temperature (27.2°C) was within tolerance [22], loss in its potency was likely through washing as has been reported elsewhere [83]. As ITS was observed intact after 4 years in the pilot Kenyan study [43], optimisation of the insecticide formulation is needed to retain insecticide content for the duration of the product’s life and to ensure cost effective control of vector borne diseases [84]. No holes were observed on the ITS after one year of installation in the animal house.

The modelling exercise performed here indicates the epidemiological potential of a house screening product that can also induce mosquito mortality. The year-old ITN performed better than the average effects we might have expected from the meta-analyses performed previously [76]. This might be due to a mis-match in our process to artificially age the ITN and what happens in reality, or it might be that the assumptions we make to estimate impacts in the SFS data for the modelling over estimate ITN performance (e.g., our assumption that mosquitoes that are repelled inside the house die). Either way, the ITS outperformed the year-old ITNs. Our like-for-like modelling comparison, however, assumes ITS is implemented overnight to the community at the 60% cover level which is likely not possible, especially as the target population increases in size. The gain from the ITS may be more gradual therefore than depicted in Figure 6.

The simulation exercise shows that the protective impact can be lasting even as mortality inducing effects wane. An advantage of this product over nets is their capacity to protect all household members equitably. While the model does not target net use to any particular group (a limitation), studies have shown that teenagers are least likely to sleep under mosquito nets [85, 86], and social hierarchies sometimes lead to vulnerable household members being exposed through no access to nets [26]. Failing to protect the sleeping space leaves individuals at risk of infection, and even if individual risks of illness are low given acquired immunity, these individuals may act as reservoirs for malaria parasite transmission [87]. The comparative analysis completed may be overestimating the initial impact given the likely longer time required to implement a house screening intervention compared to ITNs that can be deployed almost simultaneously through various operational strategies. Nevertheless, the likely durability of screening will advantage those enjoying its benefits; even while mortality impacts wane, holes are less likely to be accrued than for interventions that are manipulated more often, like ITNS. The sensitivity analysis indicates that even with relatively low cover of 40%, an impressive reduction in prevalence and cases may be achievable with a product that has a mean mortality inducing duration of only 1 to 2 years (Figure 6B, C). The modelling exercise raises the possibility of considering such a product as an alternative to ITNs, though further work is required to ensure adverse environmental impacts are minimised and that no adverse effects for users are experienced.

While the efficacy of the ITS varied by species the trends were consistent. The proportion of mortality of *An. funestus* mosquitoes was approximately 13% lower in huts installed with aged ITS than aged Olyset® Plus ITNs. This may be due to the insecticide resistance profile of the vector and also its small size enabling it to pass more easily through the ITS. However, it is important that ITS is able to kill *An. funestus* given its competence in transmission of malaria parasite [58]. The use of alternative non-pyrethroid insecticides targeting this species in ITS is warranted.

While nuisance biting vector: *Cx. quinquefasciatus* and the dengue vector *Ae. aegypti* were not killed in as high proportions as the malaria vector species despite *Ae. aegypti* being susceptible to pyrethroids (Table S1) they were still kept out of houses and prevented from blood feeding. This is extremely important because *Cx. quinquefasciatus* is a nuisance biting mosquito and protection from its bites will increase the likelihood of compliance with the ITS [88]. It is also possible that an intervention designed for malaria control may have some impact on dengue transmission if applied at scale although more work will be required to evaluate this.

Although ITS lost the majority (>70%) of their AI due to the high release rate technique, it was still able to prevent blood feeding and reduced mosquito entry into the huts installed with ITS. This is consistent with studies that have investigated house modification and insecticide treated eave nets [48, 49]. This was contributed by the physical barrier provided by the intact netting. In addition, given that there was still a little amount of AI on them, a sublethal dose of insecticide may have induce disorientation or prevent host seeking that also impact disease transmission [78, 89] and may eventually lead to mosquito mortality [90].

The risks (low mortality, high blood feeding and low deterrence) to mosquito in the huts without any treatment was extremely high, depicting the need that no house in the community should be left without any intervention. Considering that the use-life of ITNs, the conventional tool life is less than two years [91], whereas mass campaigns are often conducted every three years [10], indicates that many households are left unprotected in the community. This scenario may be the cause of stagnant or rebound of malaria prevalence in that setting, as demonstrated in Zambia [57]. Use of ITS as house modification tools are thought to be long lasting due to the low interaction with them compared to ITNs, therefore a scale up of this intervention is required so that when access to ITNs decline, and community waiting for campaigns or keep up, ITS will continue to provide protection.

There was a consistently higher number of mosquitoes recaptured in the hut without treatment. This was due to the ability of ants to gain access into the SFS. It appeared that the recapturing relied on how efficacious the treatments are in terms of mortality, in that recapture was lowest in the experimental huts with the new ITS having the highest mortality at 72 hours, indicating that mosquitoes exposure to the insecticide made them vulnerable to be eaten by predators, probably due to the knockdown or sub lethal dose [90]. This was also supported by an estimated Pearson Correlation of -0.0641 with a p-value of 0.0037 between the number of mosquitoes recaptured and number of dead mosquitoes. Ants were indeed observed in the SFS in the first round of the tests and were controlled using four ant bait stations manufactured by Neudorff, Switzerland. Each station were placed out of each chamber, therefore their presence was unlikely to have an impact on the observed efficacy of ITS, although ant control is an essential prerequisite of SFS experiments.

Perceived adverse effects, especially sneezing was reported among all the sleepers in all the huts, although this was more common with those that slept under the treated arm, indicating a response towards the exposure to the pyrethroids-PBO [22], the observed side effects are tolerable.

The SFS provides a reliable assay to measure the efficacy of this tool that requires collection of mosquitoes that have encounters the insecticidal screens to observe endpoints [89]. Bioassays are often needed to measure efficacy of vector control tools before they can be recommended or listed for public use [22, 45]. Considering the robustness on capturing important endpoints used in decision making of vector products, especially on mortality, it then makes it a feasible bioassay for assessing new house modification tools under semi-field settings and has been conducted in a number of settings [48, 92, 93]. SFS bioassays are useful because these experiments can be conducted all year round because they rely on laboratory mosquitoes of a set density, physiological status, and recapture of mosquitoes will allow reliable estimatation of the efficacy. Use of laboratory reared insects allows safe work with arbovirus vectors in areas of ongoing arbovirus transmission [94].

Although the study was relatively high throughput given the sufficient replicates and natural aging of the products and control of the bioassay. Studies for over three years would have provided information to the comparative efficacy of the product to ITNs. We recommend future studies to be conducted with ITS aged for a longer period and with slow release non-pyrethroid insecticides. In the scenario analysis, we assumed relatively high indoor biting behaviours for mosquitoes given the estimates observed in SSA [38], and this will render greater impacts than for areas where mosquitoes show greater exophilic behaviours – however this is true too for other indoor interventions [95]. Our model parameters are based on a single SFS study. The estimated parameters for the ITN tested show impacts in the upper region of the meta-analysis for ITNs [67, 74], so caution is needed given the results may be less powerful if repeated elsewhere, and it may be useful to investigate other bioassays to evidence impact. Our assumption on mosquitoes having to pass the housing barrier twice, improved the benefits from the ITS. But this seems reasonable as the barrier, and corresponding potential for mosquitoes to contact the mortality-inducing chemistries, remains in place as a mosquito tries to exit. Some may be able to leave via doors, but we have not empirically tested this here and our assumption is that this is not occurring. Empirical studies are needed for the product to allow the model to be validated if using SFS systems, or other assays, to determine and quantify the mechanism of action for modelling to be robust enough to extrapolate results more widely.

## Conclusion

Our work strongly suggests that ITS can be an efficacious tool for controlling vectors transmitting malaria, and dengue, and those known for nuisance biting in a semi-field setting. Given its simplicity, it should be considered as an additional (or stand-alone) tool alongside behavioural change educational efforts to encourage the repurposing of old ITNs for house screening. Given the tools’ simplicity, motivation for repurposing of aged ITNs in the community for house modification will provide protection against vector mosquitoes. The design of low contacts with residents also allows for exploration of higher doses of insecticide or utilisation of non-classical insecticide for insecticide maintenance.

## Data Availability

All data produced in the present study are available upon reasonable request to the authors.

## Abbreviations

Ae: Aedes
AI: Active Ingredients
*An*: Anopheles
CI: Confidence Interval
*Cx*: Culex
dh: Degree of hardness
HPLC: High Performance Liquid Chromatography
IHI: Ifakara Health Institute
IQR: Interquartile range
IRR: Incidence Rate Ratio
IRS: Indoor Residual Spraying
ITENs: Insecticide Treated Eave Nets
ITNs: Insecticide Treated Nets
ITWS: Insecticide Treated Window Screens
M: Metres
M72: Mortality at 72 hours
OR: Odds ratio
PBO: piperonyl-butoxide
PPA: Personal Protective Equipment
RH: Relative Humidity
SFS: Semi-field System
SSA: sub-Saharan Africa
UV: Ultraviolet
WHO: World Health Organization

## Contributors

OGO designed and implemented the experiment, collected and analysed data, and drafted the manuscript for publication. RJS conceived and implemented the model, and significantly contributed to the revision. SN, ABM, DK, and AN collected the data and contributed to the revision of the manuscript. JM and JLL designed the SFS system and revised the manuscript. ZMM and EM contributed to the designing of the study and revised the manuscript. RP provided logistics and revised the manuscript. RB and OS manufactured ITS, advised on the testing modality and revised the manuscript, they were not involved in data collection or analysis. JS provided substantial contribution to the revision of the manuscript. ESS conceived and implemented the model, drafted the model descriptions and significantly contributed to the revision. JB and SJM conceived the study, supervised the study and extensively contributed to the revision. All the authors approved the final draft of the manuscript for publication.

## Acknowledgements

A well-deserved appreciation to the volunteers for participating in the study and the VCPTU’s team for their hard work and contribution towards the implementation of the study. A special appreciation to Mrs Ester Mbega, and Mrs Ritha Kisava for their unflinching administrative support and Mr. Mzee Mangapi for rearing the mosquitoes used in the study. We also acknowledge the efforts of the team at Biolytrics Vietnam Co., Ltd, Hanoi, Vietnam, Vegro Aps, Copenhagen, Denmark and MCC47, Montpellier, France for developing the insecticide used in this study. We acknowledge Rob McCann from the Center for Vaccine Development and Global Health, University of Maryland School of Medicine, Baltimore, USA, for scientific input on the modelling work.

## Ethics approval and consent to participate

Participants (adults male) were enrolled from the villages at close proximity to IHI Bagamoyo, based on written informed consent. Technicians that reared mosquitoes and tested the insecticides in this study were trained and provided with necessary personal protection equipment. Volunteers were screened for malaria weekly when participating in the study. The approval to conduct the study was granted by the Ifakara Health Institute-Institutional Review Board (IHI/IRB/No: 19-2020), National Institute for Medical Research Tanzania (NIMR), Tanzania (NIMR/HQ/R.8a/Vol.IX/3473) and London School of Hygiene and Tropical Medicine (LSHTM) Observational / Interventions Research Ethics Committee (21639 – 1).

## Consent for publication

Permission to publish was sought from National Institute of Medical Research (NIMR), Tanzania.

