## Supplementary figures and images for "House modifications using insecticide treated screening of eave and window as vector control tool: evidence from a semi-field system in Tanzania and simulated epidemiological impact"

### Supplementary Figure 1

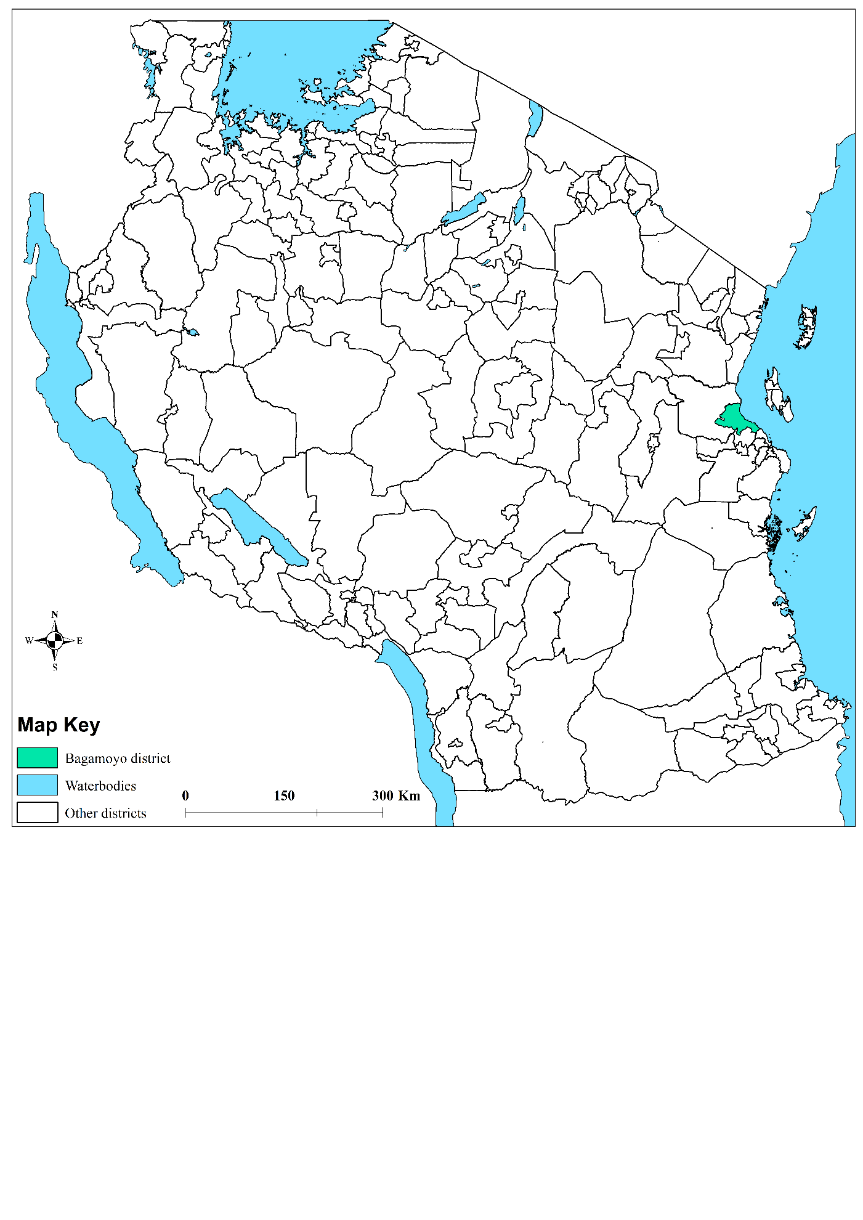

*Map generated by Yeromin P Mlacha (IHI)*

### Supplementary Figure 2

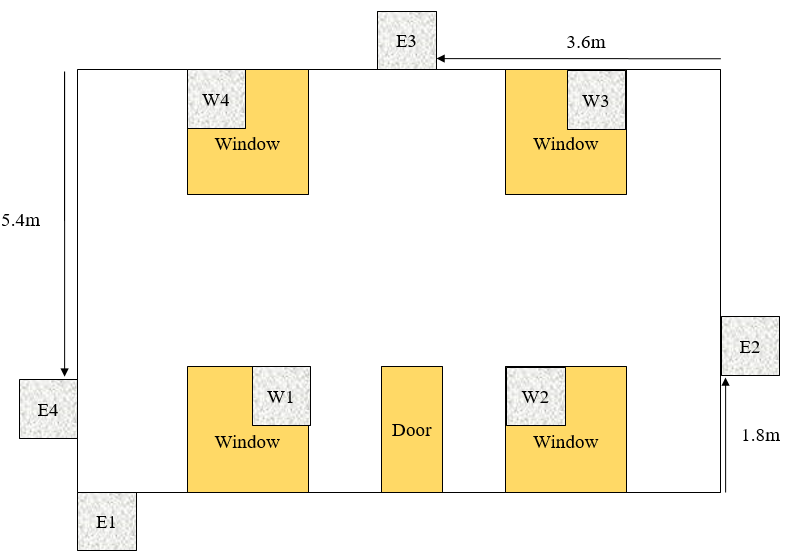

### Supplementary Figure 3

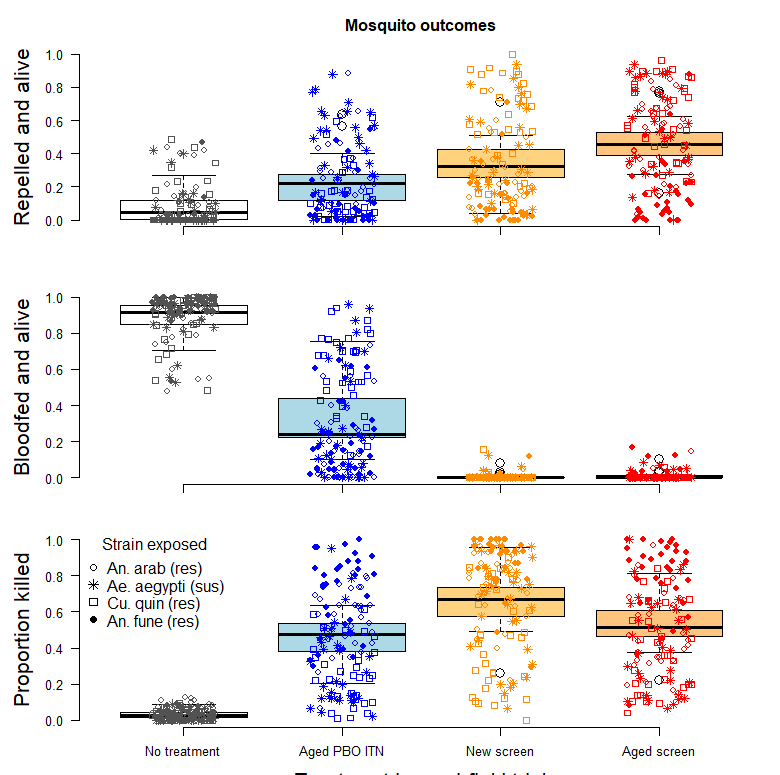
