## Supplementary Table 1 for "House modifications using insecticide treated screening of eave and window as vector control tool: evidence from a semi-field system in Tanzania and simulated epidemiological impact"

| Test systems | Origin | colonisation | established in Bagamoyo | Testing period | Permethrin (0.75%) | Deltamethrin (0.05%) | Alpha-cypermethrin (0.05%) | Lambda-cyhalothorin (0.05%) | Pirimiphos methyl (0.25%) | PBO (4%) | Permethrin (0.75%) + PBO | Deltamethrin (0.05%) + PBO | Alpha-cypermethrin (0.05%) + PBO | Lambda-cyhalothorin (0.05%) + PBO |
| --- | --- | --- | --- | --- | --- | --- | --- | --- | --- | --- | --- | --- | --- | --- |
| *An. arabiensis*  (Kingani) | Kilombero, Tanzania | 2005 | 2011 | April 2021 | 3% | 46% | 12% | 22% | 100% | 0% | 98% | 100% | 100% | 100% |
|  |  |  |  | March 2022 | 11% | 22% | 17% | 21% | 98% | 5% | 99% | 100% | 100% | 100% |
| *An. funestus*  (FUMOZ) | NICD South Africa | 2000 | 2021 | Feb 2021 | 61% | 34% | 10% | 17% | 100% | 2% | 100% | 100% | 100% | 100% |
|  |  |  |  | March 2022 | 60% | 78% | 72% | 66% | 100% | 6% | 100% | 100% | 100% | 100% |
| *Aedes aegypti*  (Bagamoyo) | Bagamoyo, Tanzania | 2015 | 2015 | Aug 2021 | 100% | 100% | 100% | 100% | 100% | - | - | - | - | - |
|  |  |  |  | March 2022 | 100% | 100% | 100% | 100% | 97% | 3% | - | - | - | - |
| *Culex quinquefasciatus*  (Bagamoyo) | Bagamoyo, Tanzania | 2019 | 2019 | March 2021 | 10% | 39% | 17% | 29% | 90% | 2% | 65% | 77% | 55% | 81% |
|  |  |  |  | March 2022 | 14% | 7% | 11% | 32% | 45% | 1% | 52% | 48% | 44% | 58% |

- Denotes that tests were not conducted at that time.
