## Supplementary Table 2 for "House modifications using insecticide treated screening of eave and window as vector control tool: evidence from a semi-field system in Tanzania and simulated epidemiological impact"

| Parameter and description | | *An. arabiensis*-like | *An. funestus*-like |
| --- | --- | --- | --- |
| *Q_0_* | Anthropophagy | 0.71 | 0.94 |
| *Φ_I_* | Proportion in mosquitoes attempting to bite humans indoors in the absence of interventions | 0.86 | 0.87 |
| *Φ_B_* | Proportion of mosquitoes attempting to bite humans in bed in the absence of interventions | 0.80 | 0.78 |
|  | Blood meal rates | 0.33 | 0.33 |
|  | Foraging time | 0.69 | 0.69 |
|  | Background mortality rate | 0.132 | 0.112 |
