## Supplementary Table 3 for "House modifications using insecticide treated screening of eave and window as vector control tool: evidence from a semi-field system in Tanzania and simulated epidemiological impact"

| Species |  | No treatment | AGED Olyset® Plus | AGED DM+PBO | NEW DM+PBO |
| --- | --- | --- | --- | --- | --- |
| **Overall (Combination of all species)** | N females released | 3990 | 3935 | 3930 | 3867 |
|  | N females recaptured | 3710 | 3209 | 3424 | 3027 |
|  | % of females recaptured (95% CI) | 92.7 (90.9 - 94.5) | 81.1 (78.1 - 84.1) | 86.9 (84.2 - 89.5) | 78.2 (74.9 - 81.4) |
| *An. arabiensis*  (Kingani strain) | N females released | 987 | 970 | 964 | 962 |
|  | N females recaptured | 898 | 805 | 848 | 749 |
|  | % of females recaptured (95% CI) | 91.0 (87.8 - 94.3) | 82.4 (77.5 - 87.3) | 88.0 (84.3 - 91.8) | 78.0 (72.2 - 83.8) |
| *An. funestus*  (FUMOZ strain) | N females released | 1012 | 986 | 972 | 970 |
|  | N females recaptured | 963 | 733 | 773 | 717 |
|  | % of females recaptured | 94.2 (90.9 - 97.4) | 72.6 (65.2 - 79.9) | 79.3 (72.7 - 85.9) | 73.7 (66.7 - 80.8) |
| *Cx quinquefasciatus* (Bagamoyo strain) | N females released | 987 | 985 | 988 | 969 |
|  | N females recaptured | 903 | 823 | 901 | 800 |
|  | % of females recaptured | 91.5 (87.3 - 95.6) | 85.2 (78.3 - 92.1) | 90.9 (87.1 - 94.7) | 83.2 (78.3 - 88.1) |
| *Ae. aegypti*  (Bagamoyo strain) | N females released | 1004 | 994 | 1006 | 966 |
|  | N females recaptured | 946 | 848 | 902 | 761 |
|  | % of females recaptured | 94.3 (91.1 - 97.4) | 84.3 (78.5 - 90.0) | 89.2 (83.7 - 94.6) | 82.1 (76.9 - 87.3) |

Shaded column: Investigational ITENs and ITWS. 95% Confidence intervals represent observations for the 32 night of experiments.
