## Supplementary Table 4 for "House modifications using insecticide treated screening of eave and window as vector control tool: evidence from a semi-field system in Tanzania and simulated epidemiological impact"

| Species | Test items | Total recaptured (N) | Total mortality (n) | %Arithmetic mean (95% CI) | Odds ratio  (95% CI) | P-value |
| --- | --- | --- | --- | --- | --- | --- |
| **Overall (All mosquitoes)** | Aged Olyset Plus | **3,209** | **1,748** | **54.9 (50.0 – 59.8)** | **1.00** |  |
|  | Aged ITS | **3,424** | **1,670** | **50.6 (45.7 – 55.5)** | **0.80 (0.59 – 1.08)** | **0.141** |
|  | New ITS | **3,027** | **2,002** | **67.6 (62.4 – 72.8)** | **2.25 (1.65-3.06)** | **<0.0001** |
|  | No treatment | **3,710** | **204** | **5.6 (4.7 – 6.5)** | **0.03 (0.02 – 0.04)** | **<0.0001** |
| *An. arabiensis*  (Kingani strain) | Aged Olyset Plus | 805 | 561 | 68.0 (60.9, 75.2) | 1.00 |  |
|  | Aged ITS | 848 | 542 | 63.9 (56.5, 71.3) | 0.82 (0.62 **–** 1.10) | 0.183 |
|  | New ITS | 749 | 609 | 81.8 (76.3, 87.3) | 2.36 (1.17 **–** 3.26) | <0.0001 |
|  | No treatment | 898 | 64 | 7.5 (5.4, 9.6) | 0.03 (0.02 **–** 0.04) | <0.0001 |
| *An. funestus*  (FUMOZ strain) | Aged Olyset Plus | 733 | 565 | 78.7 (72.0, 85.3) | 1.00 |  |
|  | Aged ITS | 773 | 500 | 65.7 (57.1, 74.3) | 0.44 (0.27 **–** 0.72) | 0.001 |
|  | New ITS | 717 | 620 | 87.9 (81.5, 94.4) | 2.41 (1.40 **–** 4.16) | 0.002 |
|  | No treatment | 963 | 52 | 5.5 (3.7, 7.3) | 0.01 (0.00 **–** 0.01) | <0.0001 |
| *Cx. quinquefasciatus* (Bagamoyo strain) | Aged Olyset Plus | 823 | 308 | 36.1 (28.7, 43.5) | 1.00 |  |
|  | Aged ITS | 901 | 323 | 36.2 (26.6, 45.8) | 0.97 (0.65 **–** 1.43) | 0.859 |
|  | New ITS | 800 | 341 | 43.3 (33.3, 53.3) | 1.39 (0.94 **–** 2.06) | 0.103 |
|  | No treatment | 903 | 47 | 5.2 (3.5, 6.9) | 0.07 (0.04 **–** 0.12) | <0.0001 |
| *Ae. aegypti*  (Bagamoyo strain) | Aged Olyset Plus | 848 | 314 | 36.8 (28.7, 44.9) | 1.00 |  |
|  | Aged ITS | 902 | 305 | 36.7 (28.1, 45.2) | 0.98 (0.60 **–** 1.61) | 0.950 |
|  | New ITS | 761 | 432 | 57.5 (47.2, 67.7) | 3.01 (1.82 **–** 4.96) | <0.0001 |
|  | No treatment | 946 | 41 | 4.3 (2.8, 5.8) | 0.06 (0.03 **–** 0.11) | <0.0001 |

Treatment, volunteer, hut and day are fixed effect and observation as random effect. For overall, species was included as a fixed effect.
