## Supplementary Table 5 for "House modifications using insecticide treated screening of eave and window as vector control tool: evidence from a semi-field system in Tanzania and simulated epidemiological impact"

| Species | Test items | Total recaptured (N) | Total blood feeding (n) | %Arithmetic mean (95% CI) | Odds ratio (95% CI) | P-value |
| --- | --- | --- | --- | --- | --- | --- |
| **Overall (Combination of all mosquitoes)** | **Aged Olyset Plus** | **3,209** | **1,537** | **46.3 (40.7 – 51.9)** | **1.00** |  |
|  | **Aged ITS** | **3,424** | **511** | **15.5 (12.0 – 19.0)** | **0.09 (0.05 – 0.14)** | **<0.0001** |
|  | **New ITS** | **3,027** | **178** | **6.3 (3.7 – 9.0)** | **0.02 (0.01 – 0.03)** | **<0.0001** |
|  | ***No treatment*** | **3,710** | **3,447** | **92.8 (90.6 – 95.0)** | **66.33 (39.50 – 111.38)** | **<0.0001** |
| *An. arabiensis*  (Kingani strain) | Aged Olyset Plus | 805 | 281 | 35.0 (25.2 – 44.9) | 1.00 |  |
|  | Aged ITS | 848 | 33 | 4.5 (0.2 – 8.7) | 0.02 (0.01 – 0.05) | <0.0001 |
|  | New ITS | 749 | 3 | 0.4 (0 – 0.9) | 0.00 (0.00 – 0.01) | <0.0001 |
|  | *No treatment* | 898 | 813 | 90.2 (85.3 – 95.2) | 75.16 (32.38 – 174.42) | <0.0001 |
| *An. funestus*  (FUMOZ strain) | Aged Olyset Plus | 733 | 352 | 42.4 (32.2 – 52.6) | 1.00 |  |
|  | Aged ITS | 773 | 264 | 34.4 (25.3 – 43.6) | 0.63 (0.32 – 1.24) | 0.182 |
|  | New ITS | 717 | 58 | 7.7 (2.2 – 13.2) | 0.04 (0.02 – 0.08) | <0.0001 |
|  | *No treatment* | 963 | 942 | 97.7 (94.7 – 100) | 335.91 (116.87 – 965.50) | <0.0001 |
| *Cx quinquefasciatus* (Bagamoyo strain) | Aged Olyset Plus | 823 | 597 | 71.2 (62.8 – 79.7) | 1.00 |  |
|  | Aged ITS | 901 | 115 | 12.5 (8.0 – 17.0) | 0.03 (0.01 – 0.05) | <0.0001 |
|  | New ITS | 800 | 63 | 7.6 (1.9 – 13.4) | 0.01 (0.01 – 0.02) | <0.0001 |
|  | *No treatment* | 903 | 811 | 90.8 (85.9 – 95.7) | 7.18 (3.77 – 13.66) | <0.0001 |
| *Ae. aegypti*  (Bagamoyo strain) | Aged Olyset Plus | 848 | 307 | 36.6 (25.0 – 48.1) | 1.00 |  |
|  | Aged ITS | 902 | 99 | 10.7 (6.8 – 14.6) | 0.14 (0.06 – 0.31) | <0.0001 |
|  | New ITS | 761 | 54 | 9.6 (2.7 – 16.6) | 0.07 (0.03 – 0.17) | <0.0001 |
|  | *No treatment* | 946 | 881 | 92.6 (88.2 – 96.9) | 95.04 (39.37 – 229.44) | <0.0001 |

Treatment, volunteer, hut and day are fixed effect and observation as random effect. For overall, specie was included as a fixed effect.
