## Supplementary Table 6 for "House modifications using insecticide treated screening of eave and window as vector control tool: evidence from a semi-field system in Tanzania and simulated epidemiological impact"

| Species |  | Total recaptured (N) | Total indoor (n) | Median (IQR) | IRR (95% CI) | P-value |
| --- | --- | --- | --- | --- | --- | --- |
| **Overall (All study strain)** | **Aged Olyset Plus** | **3,209** | **1,832** | **13 (8 – 22)** | **1.00** |  |
|  | **Aged ITS** | **3,424** | **583** | **2 (0 – 6)** | **0.25 (0.21 – 0.31)** | <0.0001 |
|  | **New ITS** | **3,027** | **271** | **0 (0 – 2)** | **0.10 (0.08 – 0.13)** | <0.0001 |
|  | ***No treatment*** | **3,710** | **3,022** | **25 (20 – 29)** | **1.73 (1.46 – 2.06)** | <0.0001 |
| *An. arabiensis*  (Kingani strain) | Aged Olyset Plus | 805 | 344 | 10 (8 – 12) | 1.00 |  |
|  | Aged ITS | 848 | 43 | 0 (0 – 1) | 0.09 (0.05, 0.14) | <0.0001 |
|  | New ITS | 749 | 39 | 0 | 0.05 (0.03, 0.09) | <0.0001 |
|  | *No treatment* | 898 | 688 | 21 (18 – 27) | 1.98 (1.42, 2.75) | <0.0001 |
| *An. funestus*  (FUMOZ strain) | Aged Olyset Plus | 733 | 384 | 10 (5 – 20) | 1.00 |  |
|  | Aged ITS | 773 | 248 | 6 (2 – 12) | 0.57 (0.40, 0.79) | 0.001 |
|  | New ITS | 717 | 66 | 0 (0 – 2) | 0.12 (0.08, 0.19) | <0.0001 |
|  | *No treatment* | 963 | 817 | 28 (22 – 31) | 2.45 (1.79, 3.35) | <0.0001 |
| *Cx quinquefasciatus* (Bagamoyo strain) | Aged Olyset Plus | 823 | 599 | 20 (14 – 25) | 1.00 |  |
|  | Aged ITS | 901 | 152 | 3 (1 – 8) | 0.21 (0.15, 0.29) | <0.0001 |
|  | New ITS | 800 | 80 | 1 (0 – 2) | 0.08 (0.05, 0.12) | <0.0001 |
|  | *No treatment* | 903 | 771 | 27 (21 – 29) | 1.32 (1.01, 1.74) | 0.043 |
| *Ae. aegypti*  (Bagamoyo strain) | Aged Olyset Plus | 848 | 505 | 17 (8 – 22) | 1.00 |  |
|  | Aged ITS | 902 | 140 | 3 (1 – 5) | 0.20 (0.15, 0.28) | <0.0001 |
|  | New ITS | 761 | 86 | 1 (0 – 3) | 0.16 (0.12, 0.23) | <0.0001 |
|  | *No treatment* | 946 | 746 | 24 (18 – 29) | 1.59 (1.21, 2.08) | 0.001 |

Treatment, volunteer, hut and day are fixed effect and observation as random effect. For overall, specie was included as a fixed effect.
