## Supplementary Table 7 for "House modifications using insecticide treated screening of eave and window as vector control tool: evidence from a semi-field system in Tanzania and simulated epidemiological impact"

| Type of Adverse Event | NO TREATMENT | AGED OLYSET® PLUS ITNs | AGED ITS | NEW ITS |
| --- | --- | --- | --- | --- |
|  | N | N | N | N |
| Skin Itching | 0 | 0 | 0 | 0 |
| Facial Burning | 0 | 0 | 0 | 1 |
| Sneezing | 1 | 1 | 3 | 3 |
| Running Nose | 0 | 0 | 0 | 0 |
| Headache | 0 | 0 | 0 | 1 |
| Nausea | 0 | 0 | 0 | 0 |
| Eye Irritation | 1 | 0 | 2 | 0 |
| Tears from eyes | 0 | 0 | 0 | 0 |
| Bad Smell | 0 | 0 | 0 | 1 |
| TOTAL | **2** | **1** | **5** | **6** |

The total observation is 16, that is 4 people per night for the first 4 nights of the experiment
