## Supplementary Table 8 for "House modifications using insecticide treated screening of eave and window as vector control tool: evidence from a semi-field system in Tanzania and simulated epidemiological impact"

|  |  | **Quality checks before trial** | | | **Quality checks after trial** | | | |
| --- | --- | --- | --- | --- | --- | --- | --- | --- |
| **TEST SYSTEM** | **TEST ITEM** | **No. of mosquitoes** | **% KD60**  **(95% CI)** | **% 24hr control corrected Mortality**  **(95% CI)** | **Test item** | **No. of mosquitoes** | **% KD60**  **(95% CI)** | **% 72hr control corrected Mortality**  **(95% CI)** |
| Overall (Combination of all mosquitoes) | New Olyset Plus | 180 | 100 | 93.3  (89.1 – 97.5) | Aged Olyset Plus | 400 | 7.0  (4.7 – 9.3) | 15.1  (9.4 – 20.7) |
|  | New ITENs & ITWS | 240 | 77.9  (66.7 – 89.2) | 67.1  (55.4 – 78.7) | Aged ITENs and ITWS | 640 | 15.9  (11.9 – 20.0) | 10.2  (6.7 – 13.7) |
|  |  |  |  |  | New ITENs and ITWS | 640 | 38.1  (31.2 – 45.1) | 22.7  (17.1 – 28.2) |
| Pyrethroid-resistant *An. arabiensis*  (Kingani) | New Olyset Plus | 60 | 100 | 100 | Aged Olyset Plus | 100 | 9.0  (3.7 – 14.3) | 22.0  (11.0 – 33.0) |
|  | New ITENs & ITWS | 60 | 100 | 96.7  (92.2 – 100) | Aged ITENs and ITWS | 160 | 26.9  (18.9 – 34.9) | 21.5  (11.0 – 32.0) |
|  |  |  |  |  | New ITENs and ITWS | 160 | 60.0  (49.5 – 70.5) | 16.9  (10.5 – 23.3) |
| Pyrethroid-resistant *An. funestus (FUMOZ)* | New Olyset Plus | - | - | - | Aged Olyset Plus | 100 | 4.0  (0.4 – 7.6) | 4.0  (0.4 – 7.6) |
|  | New ITENs & ITWS | 60 | 100 | 73.3  (60.1 – 86.6) | Aged ITENs and ITWS | 160 | 0.6  (0 – 1.9) | 4.4  (0.5 – 8.2) |
|  |  |  |  |  | New ITENs and ITWS | 160 | 4.4  (1.0 – 7.8) | 6.9  (1.4 – 12.4) |
| Pyrethroid-susceptible *Ae. aegypti* (Bagamoyo) | New Olyset Plus | 60 | 100 | 100 | Aged Olyset Plus | 100 | 13.0  (7.8 – 18.2) | 24.2  (10.0 – 38.4) |
|  | New ITENs & ITWS | 60 | 100 | 95.0  (89.8 – 100) | Aged ITENs and ITWS | 160 | 33.1  (23.2 – 43.1) | 13.1  (6.9 – 19.4) |
|  |  |  |  |  | New ITENs and ITWS | 160 | 83.1  (75.0 – 91.2) | 65.0  (54.1 – 75.9) |
| Pyrethroid-resistant *Cx. quinquefasciatus* (Bagamoyo) | New Olyset Plus | 60 | 100 | 80.0  (71.5 – 88.5) | Aged Olyset Plus | 100 | 2.0  (0 – 4.7) | 10.0  (0 – 21.6) |
|  | New ITENs & ITWS | 60 | 11.7  (4.0 – 19.3) | 3.3  (0 – 7.8) | Aged ITENs and ITWS | 160 | 3.1  (0.6 – 5.7) | 1.9  (0 – 3.9) |
|  |  |  |  |  | New ITENs and ITWS | 160 | 5.0  (1.5 – 8.5) | 1.9  (0, 3.9) |

Dash indicates that test was not conducted.
