## Supplementary Table 9 for "House modifications using insecticide treated screening of eave and window as vector control tool: evidence from a semi-field system in Tanzania and simulated epidemiological impact"

| **Content** | **Number** | **AI content (g/kg)**  **Arithmetic Mean (SD)** | | **% Within ±25 target dose** | | **% AI content retained after aging** |
| --- | --- | --- | --- | --- | --- | --- |
|  |  | **NEW ITS** | **AGED ITS** | **NEW ITS** | **AGED ITS** | **ITS** |
| Deltamethrin | 4 | 2.5 (0.1) | 0.7 (0.4) | 100 | 0 | 27.0 |
| PBO | 4 | 10.2 (0.1) | 0.9 (0.4) | 100 | 0 | 8.4 |

***Estimates were calculated using estimates from individual pieces of net to account for variation within net.***
